# Comparative effectiveness of natalizumab and fingolimod and injectable therapies in patients with pediatric multiple sclerosis: A registry-based retrospective cohort study

**DOI:** 10.1101/2022.10.12.22280969

**Authors:** Tim Spelman, Gabrielle Simoneau, Robert Hyde, Robert Kuhelj, Raed Alroughani, Serkan Ozakbas, Rana Karabudak, Bassem Yamout, Samia J. Khoury, Murat Terzi, Cavit Boz, Dana Horakova, Eva Kubala Havrdova, Bianca Weinstock-Guttman, Francesco Patti, Ayse Altintas, Saloua Mrabet, Jihad Inshasi, Helmut Butzkueven, the MSBase Investigators

## Abstract

**Background and Objectives:** Patients with pediatric-onset multiple sclerosis (POMS) typically experience higher levels of inflammation with more frequent relapses and reach irreversible disability at a younger age than adult-onset patients. There have been few randomized placebo-controlled clinical trials of multiple sclerosis (MS) disease-modifying therapies (DMTs) in patients with POMS, and most available data are based on observational studies of off-label use of DMTs approved for adults. We assessed the effectiveness of natalizumab compared with fingolimod using injectable platform therapies as a reference in pediatric patients in the global MSBase registry.

**Methods:** This retrospective study included patients with POMS who initiated treatment with an injectable DMT, natalizumab, or fingolimod between January 1, 2006, and May 3, 2021 (N=1218). The primary outcome was the time to first relapse from index therapy initiation. Secondary study outcomes included annualized relapse rate; proportions of relapse-free patients at 1, 2, and 5 years post baseline; time to treatment discontinuation; and times to 24-week confirmed disability worsening and confirmed disability improvement.

**Results:** Patients treated with fingolimod had a significantly lower risk of relapse than patients treated with injectable DMT (hazard ratio [HR], 0.49; 95% confidence interval [CI], 0.29–0.83; *P*=0.008). After adjustment for prior DMT experience in the unmatched sample, patients treated with natalizumab had a significantly lower risk of relapse than patients treated either with injectable DMT (HR, 0.15; 95% CI, 0.07–0.31; *P*<0.001) or fingolimod (HR, 0.37; 95% CI, 0.14–1.00; *P*=0.049). The adjusted secondary study outcomes were generally consistent with the primary outcome or with previous observations. The findings in the inverse probability treatment weighting–adjusted patient populations were confirmed in multiple sensitivity analyses.

**Discussion:** Our results suggest that natalizumab and fingolimod have broadly equivalent therapeutic efficacies in patients with POMS, consistent with previous studies of natalizumab and fingolimod in adult-onset patients and POMS. However, analyses of relapse outcomes suggest natalizumab is superior to fingolimod in the control of relapses in this population with high rates of new inflammatory activity.

**Classification of Evidence:** This study provides Class III evidence that natalizumab may provide better disease control than fingolimod in patients with POMS.

## INTRODUCTION

Multiple sclerosis (MS) is a chronic inflammatory demyelinating and neurodegenerative disease of the central nervous system, with an estimated worldwide prevalence in 2020 of more than 2.8 million cases.^1^ Although symptoms of MS usually first appear in adults between 20 and 50 years of age,^2^ approximately 3%–5% of MS cases are of pediatric onset, with first symptoms in childhood or, more commonly, in adolescence.^3-5^

Patients with pediatric-onset MS (POMS) typically experience higher levels of inflammation with more frequent relapses than patients with adult-onset MS (AOMS).^6-8^ Although pediatric patients take longer than adults to reach irreversible disability, this point still occurs at a younger age.^9^ A study of 394 patients with POMS found that patients exhibited a median time of 28.9 years to reach an Expanded Disability Status Scale (EDSS) score of 6, when patients were a median age of 42.2 years old, approximately 10 years younger than the average age to EDSS 6 for AOMS.^9^ Additionally, patients with POMS generally have poorer cognitive performance and long-term socioeconomic outcomes than do patients diagnosed with AOMS or healthy controls, probably because brain inflammation can interfere with ongoing cerebral maturation processes in adolescence.^10,11^ Early intervention with an appropriate efficacious disease-modifying therapy (DMT) is therefore essential for reducing the risk of persistent long-term disability in these patients.^12^

There have been few randomized placebo-controlled clinical trials of MS DMTs in pediatric patients with POMS, and therefore many of the available data are based on observational studies of off-label use of DMTs approved for adults.^13^ Patients with pediatric- and adult-onset MS share similar genetic and environmental risk factors, suggesting similar pathophysiology, and therefore pediatric patients typically show similar responses to DMTs.^14,15^ The most widely used first-line therapies in POMS patients have been injectable DMTs, including interferon-β and glatiramer acetate.^16^ However, these first-line agents may be poorly tolerated or fail to provide adequate control of disease activity,^17^ prompting the need to escalate therapy to more efficacious DMTs.^18^

Fingolimod was approved in the United States and European Union for pediatric patients with MS in 2018, on the basis of the randomized clinical trial of fingolimod versus interferon beta-1a in pediatric patients with MS.^19-21^ Observational studies of pediatric patients with MS have reported effectiveness of other DMTs as well, including dimethyl fumarate and natalizumab, though the latter has predominantly been studied as a second-line therapy in patients with breakthrough disease.^13,22-30^

Comparative effectiveness data in POMS patients are also limited. In the absence of comparative data from head-to-head randomized clinical trials, observational studies can provide useful information for treatment decision-making. We therefore assessed the effectiveness of natalizumab compared with fingolimod and with reference to the injectable platform therapies (subcutaneous [SC] interferon beta-1b, intramuscular [IM] or SC interferon beta-1a, IM or SC peginterferon beta-1a, or SC glatiramer acetate) in patients with POMS in the global MSBase registry.^31^

## METHODS

### Data

The MSBase registry consists of anonymized patient-level data from contributing member sites.^32^ Institutional review board and ethics committee approvals are required for initiation of each site according to applicable local laws and regulations. Written informed consent is obtained for each patient prior to their inclusion in the database in accordance with the Declaration of Helsinki. Data were collected from 2006, when natalizumab first became available, through 2021. A sensitivity analysis was performed using data as of 2010, when fingolimod became available for use.

### Study design and sample

This was a retrospective cohort study based on MSBase registry data from 100 centers and 32 countries. Patients diagnosed with POMS and who initiated treatment with an injectable DMT, natalizumab, or fingolimod between January 1, 2006, and May 3, 2021, were included so as to have a contemporary sample since the availability of the second-generation of MS treatments.

### Inclusion and exclusion criteria

Eligible patients had a diagnosis of pediatric-onset relapsing MS (RMS) and initiated index therapy treatment prior to the age of 18. Patients had to be naïve to prior therapy or to have switched from an injectable DMT. Patients who initiated treatment with an injectable DMT and who switched to either natalizumab or fingolimod during the follow-up period were assigned to the natalizumab or fingolimod cohort, respectively. Exclusion criteria included prior treatment with natalizumab, fingolimod, cladribine, rituximab, ocrelizumab, alemtuzumab, dimethyl fumarate, or teriflunomide, or prior treatment with a recognized immunosuppressive agent, such as azathioprine, methotrexate, cyclophosphamide, or mitoxantrone.

### Patient cohorts

Patients were assigned to 1 of 3 cohorts based on their index therapy: injectable DMT, natalizumab, or fingolimod. Patients contributed to a cohort from baseline (therapy initiation) until the patient discontinued the therapy or until the end of the follow-up.

### Outcomes and Assessments

#### Primary outcome

The primary outcome was the time to first relapse from index therapy initiation.

#### Secondary outcomes

Secondary study outcomes included annualized relapse rate (ARR); proportions of patients who were relapse-free at 1, 2, and 5 years post-baseline; time to treatment discontinuation; and times to 24-week confirmed disability worsening (CDW) and 24-week confirmed disability improvement (CDI) in EDSS scores. ARR was calculated during the entire duration of follow-up. Relapses were defined as new or recurrent neurologic symptoms occurring >30 days following the onset of a previous relapse. Symptoms occurring ≤30 days following a previous relapse were counted only as 1 relapse, and the onset date used in the analysis was the onset date of the first relapse. Time to treatment discontinuation was calculated during the entire duration of follow-up time. Time to 24-week CDW or 24-week CDI was calculated from baseline until disease progression or improvement, respectively. CDW was defined as a confirmed increase of ≥0.5 point in EDSS score for patients with a baseline EDSS score >5.5; ≥1.0 point for those with a baseline EDSS score between 1.0 and 5.5, inclusive; and ≥1.5 points for those with a baseline EDSS score of 0. For the confirmation of disability worsening, EDSS scores recorded within 30 days after the onset of a relapse were excluded. Initial and confirmatory disability progression had to be assessed on consecutive visits. CDI was defined as a confirmed decrease in EDSS score of ≥1 point for patients with a baseline EDSS score ≥2.0. CDI was not calculated for patients with a baseline EDSS score <2.0. For the confirmation of disability improvement, EDSS scores recorded within 30 days after the onset of a relapse were excluded. Initial and confirmatory disability improvement had to be assessed on consecutive visits.

### Statistical analyses

#### Analysis populations

All patients fulfilling all the inclusion criteria and not meeting any of the exclusion criteria were included in the analysis population. A multinomial logistic regression model was used to calculate propensity scores and inverse probability of treatment weighting (PS-IPTW) was used to balance groups by baseline patient characteristics (age, sex, disease duration, baseline EDSS, country, prior DMT, relapse count in the past year and past 2 years, magnetic resonance imaging [MRI] lesion count, and presence of gadolinium-enhanced [Gd+] lesions). Index calendar year was not included as a covariate to avoid positivity violation issues during modeling. This enabled a broader analysis that included patients enrolled prior to the availability of some treatments in their country.

#### Outcome assessments

For the primary and secondary outcome assessments, average treatment effect weights—which may be interpreted as targeting a combined population of patients treated with natalizumab, fingolimod, or injectable platform therapies—for each of the 3 treatment cohorts were estimated. Outcomes were assessed separately for pairwise treatment comparison. For the primary outcome, the adjusted cumulative probability of relapse post-baseline was estimated using a weighted Cox proportional hazard model controlling for treatment arm, with adjustment for previous DMT use (naïve vs experienced), count of prebaseline DMTs, and index year. The secondary ARR endpoint was estimated using a weighted negative binomial regression model controlling for treatment arm and with an offset for log-transformed follow-up time.

Proportions of relapse-free patients were evaluated using a weighted logistic regression model controlling for treatment arm. Secondary time-to-event analyses were performed using a weighted Cox proportional hazard model controlling for treatment arm. Errors (95% confidence intervals [CIs]) for all primary and secondary outcome analyses were calculated using robust sandwich estimation.

### Sensitivity analyses

Five sensitivity analyses were conducted to assess the validity of assumptions and robustness of the results (Supplementary Methods). For the propensity score–matching analysis, patients were matched 1:1 with replacement using propensity scores instead of IPTW. A weight-trimming analysis was performed to assess the impact of removing outlier propensity score weights by removing the first quartile of scores from the natalizumab-, fingolimod-, and injectable DMT–treated groups. For the alternative weighting analyses, the outcome analyses were repeated using average treatment effect among the treated (ATT) weighting to produce different target populations for each treatment group or the combined (overlap) patient population. A cohort entry date analysis was performed by restricting data to patients who were treated after January 1, 2010, when fingolimod was available for use. For the MRI lesion analysis, data were restricted to patients with a baseline MRI and included number of MRI lesions and presence of Gd+ lesions as covariates in the propensity score model.

A sensitivity analysis was also conducted to assess the effect of index calendar year on outcomes.

#### Descriptive statistics

Continuous variables were assessed using mean, standard deviation, median, 25th and 75th percentiles, or minimum and maximum, as appropriate. Categorical variables were summarized as frequencies and percentage. Descriptive statistics were tabulated for all baseline characteristics by cohort. The number of visits with a EDSS measurement after baseline were summarized separately by cohort as both a categorical and continuous measure. The inter-visit time was calculated as the time in months between consecutive visits with a EDSS measurement. The reasons for treatment discontinuation was reported in a descriptive manner. No adjustments for multiplicity were conducted, because the number of tests performed were determined to be small enough as to not appreciably affect the findings.

#### Missing data

In general, missing values were not imputed. Patients with missing data in any variable required for a given analysis were not included in that analysis. Tests were performed to determine whether these exclusions caused a bias. Partial dates were imputed for dates of relapse events, EDSS measurements, and therapy initiation or discontinuation.

Unknown days and months were imputed as the 1st and January, respectively.

### Data Availability

The clinical data for this study were obtained under a license agreement with MSBase (http://www.msbase.org). However, no patient-level data were disclosed as part of the study. Therefore, all data relevant to the study are presented in this manuscript and the Supplementary Materials.

## RESULTS

### Patients

As of May 3, 2021, there were 76,152 patients included in the MSBase registry, of whom 5410 were diagnosed with POMS. Of these, 1218 had RMS; initiated treatment with an injectable DMT, fingolimod, or natalizumab on or after January 1, 2006; and had a baseline EDSS assessment within 6 months of the index date (Table 1). Although baseline age, sex, and EDSS score were generally similar between groups, prior to IPTW adjustment, 50% of covariates displayed standardized differences >0.20 (Table 1). Following propensity score–based IPTW, mean standardized differences between treatment groups were <0.20 for 18 of 24 covariates (75%).

**Table 1.** Baseline characteristics for patients with pediatric-onset RMS

| Characteristic | Natalizumab<sup>a</sup><br>(n=111) | Fingolimod<sup>b</sup><br>(n=104) | Injectable DMT<sup>c</sup><br>(n=1003) | Standardized difference, unweighted |  |  | Standardized difference, weighted |  |  |
| --- | --- | --- | --- | --- | --- | --- | --- | --- | --- |
|  |  |  |  | Natalizumab<br>vs<br>fingolimod | Natalizumab<br>vs<br>injectables | Fingolimod<br>vs<br>injectables | Natalizumab<br>vs<br>fingolimod | Natalizumab<br>vs<br>injectables | Fingolimod<br>vs<br>injectables |
| Age, mean (SD), years | 15.80 (2.28) | 16.00 (2.89) | 16.06 (1.98) | 0.000 | 0.000 | -0.027 | 0.000 | 0.008 | -0.008 |
| Female, n (%) | 83 (74.8) | 75 (72.1) | 721 (71.9) | 0.060 | 0.065 | 0.005 | 0.047 | 0.063 | 0.049 |
| Country, n (%) |  |  |  |  |  |  |  |  |  |
| Australia | 14 (12.6) | 21 (20.2) | 63 (6.3) | 0.214 | -0.166 | -0.279 | 0.194 | -0.071 | -0.250 |
| Italy | 10 (9.0) | 2 (1.9) | 68 (6.8) |  |  |  |  |  |  |
| Kuwait | 32 (28.8) | 11 (10.6) | 74 (7.4) |  |  |  |  |  |  |
| Spain | 9 (8.1) | 14 (13.5) | 51 (5.1) |  |  |  |  |  |  |
| Turkey | 5 (4.5) | 25 (24.0) | 282 (28.1) |  |  |  |  |  |  |
| Czech Republic | 5 (4.5) | 2 (1.9) | 39 (3.9) |  |  |  |  |  |  |
| Iran | 0 (0.0) | 0 (0.0) | 46 (4.6) |  |  |  |  |  |  |
| Belgium | 3 (2.7) | 3 (2.9) | 29 (2.9) |  |  |  |  |  |  |
| Canada | 5 (4.5) | 1 (1.0) | 24 (2.4) |  |  |  |  |  |  |
| Other | 28 (25.2) | 25 (24.0) | 327 (32.6) |  |  |  |  |  |  |
| MS duration, mean (SD), years | 1.74 (1.71) | 1.91 (1.75) | 1.32 (1.65) | -0.096 | 0.248 | 0.343 | -0.016 | 0.031 | 0.148 |
| BL EDSS score,<sup>d</sup> median (IQR) | 1.5 (1, 2.5) | 1 (0, 2) | 1.5 (1, 2.5) | 0.292 | 0.130 | -0.161 | 0.155 | 0.063 | -0.137 |
| Prior DMT use, n (%) |  |  |  |  |  |  |  |  |  |
| Naïve | 64 (57.7) | 48 (46.2) | 865 (86.2) | 0.231 | -0.669 | -0.932 | 0.135 | -0.265 | -0.420 |
| Experienced | 47 (42.3) | 56 (53.9) | 138 (13.8) |  |  |  |  |  |  |
| Number of prior DMTs, mean (SD) | 0.60 (0.83) | 0.69 (0.80) | 0.15 (0.39) | -0.108 | 0.699 | 0.864 | -0.009 | 0.216 | 0.341 |
| Relapses in year prior to BL, mean (SD) | 1.53 (1.23) | 1.12 (1.17) | 1.07 (0.95) | 0.346 | 0.420 | 0.044 | 0.083 | 0.037 | -0.146 |
| Index year, n (%) |  |  |  |  |  |  |  |  |  |
| 2006–2010 | 22 (19.8) | 1 (1.0) | 359 (35.8) | -0.397 | 0.421 | 0.881 | 0.076 | 0.514 | 0.519 |
| 2011–2015 | 49 (44.1) | 59 (56.7) | 433 (43.2) |  |  |  |  |  |  |
| 2016+ | 40 (36.0) | 44 (42.3) | 211 (21.0) |  |  |  |  |  |  |
| Follow-up time, years<sup>e</sup> |  |  |  |  |  |  |  |  |  |
| Mean (SD) | 3.91 (2.94) | 3.03 (2.79) | 2.19 (2.45) | — | — | — | — | — | — |
| Median (IQR) | 3.21 (1.59, 5.55) | 2.38 (0.59, 4.73) | 1.37 (0.22, 3.41) | — | — | — | — | — | — |

<sup>a</sup>Patients did not have prior natalizumab treatment. <sup>b</sup>Patients did not have prior fingolimod treatment. <sup>c</sup>Includes IM or SC interferon beta-1a, SC interferon beta-1b, SC glatiramer acetate, and IM or SC peginterferon beta-1a. <sup>d</sup>Nearest EDSS score within 6 months of BL. <sup>e</sup>Follow-up time while on index DMT.
BL, baseline; DMT, disease-modifying therapy; EDSS, Expanded Disability Status Scale; IM, intramuscular; IQR, interquartile range; MS, multiple sclerosis; RMS, relapsing multiple sclerosis; SC, subcutaneous; SD, standard deviation.

Patients who were prescribed injectable DMTs as the index therapy had shorter MS disease duration and fewer relapses in the past year than did those who received natalizumab or fingolimod.

### Outcomes

#### Time to first relapse

Kaplan-Meier estimated proportions of relapse-free patients at years 1, 2, and 5 were greatest among those treated with natalizumab and smallest among patients treated with an injectable DMT (Figure 1). At year 1, the proportion of relapse-free patients (95% CI) was highest for natalizumab (94.8% [86.6–98.0]), followed by fingolimod (88.2% [76.9–94.2]) and injectable DMT (73.3% [69.8–76.6]). At year 2, the proportion of relapse-free patients (95% CI) was highest for natalizumab (93.4% [84.8–97.1]), followed by fingolimod (82.9% [70.4–90.4]) and injectable DMT (59.8% [55.6–63.7]). At year 5, the proportion of relapse-free patients (95% CI) was highest for natalizumab (90.0% [80.1–95.1]), followed by fingolimod (71.9% [55.5–83.1]) and injectable DMT (35.8% [30.6–40.9]).

**Figure 1.**
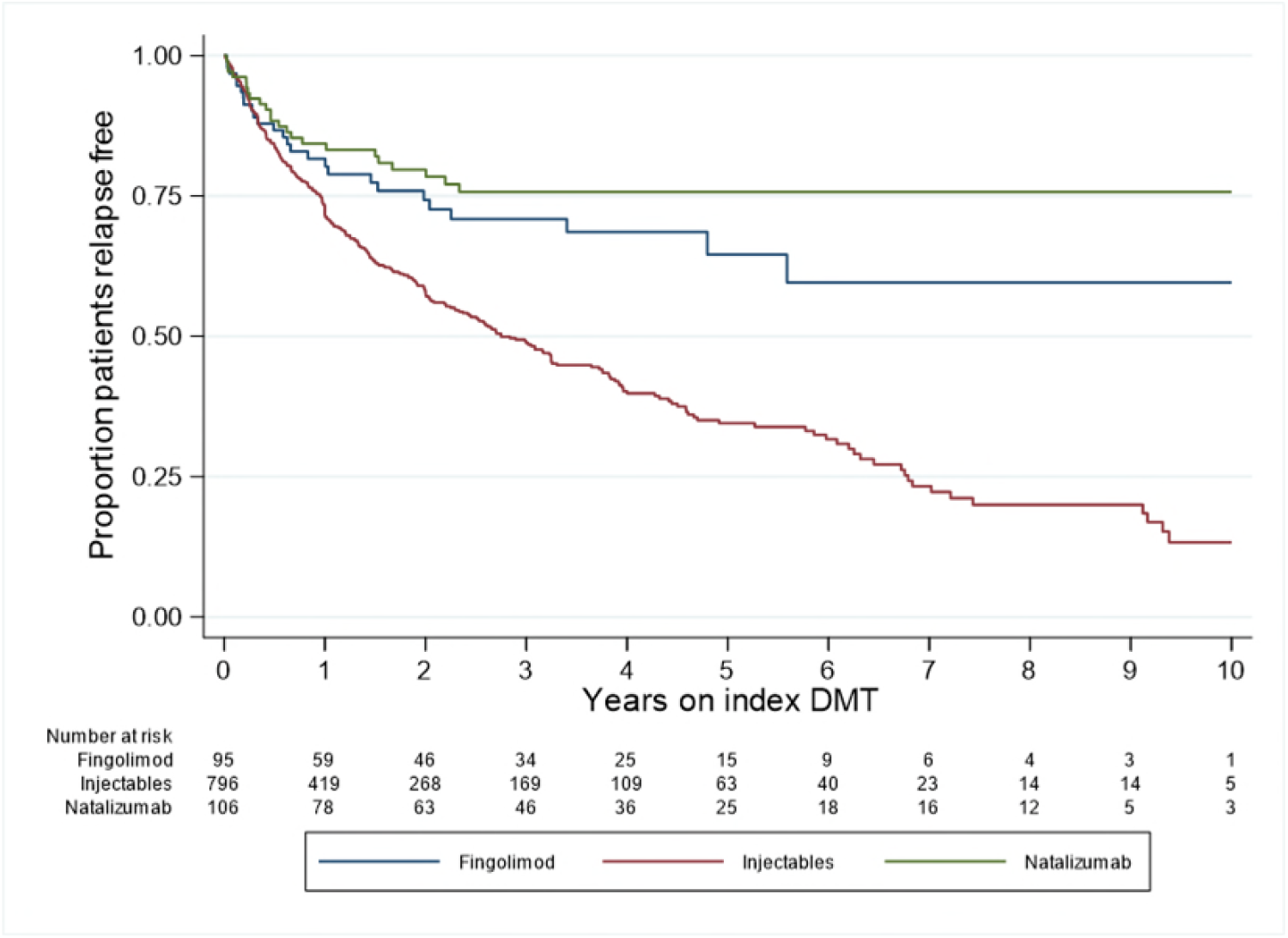
Cumulative probabilities of remaining relapse-free for patients treated with natalizumab, fingolimod, or injectable therapies. DMT, disease-modifying therapy.

Patients treated with natalizumab demonstrated significantly lower risk of relapse than did patients treated with an injectable DMT (adjusted HR, 0.15; 95% CI, 0.07–0.31; *P*<0.001) and a lower risk of relapse than patients treated with fingolimod (adjusted HR, 0.37; 95% CI, 0.14–1.00; *P*=0.049) (Table 2). Patients treated with fingolimod also had a significantly lower risk of relapse than those treated with injectable DMTs (adjusted HR, 0.49; 95% CI, 0.29–0.83; *P*=0.008).

**Table 2.** PS-IPTW adjusted Kaplan-Meier estimates of relative risk of relapse

| Comparison | PS-IPTW adjusted HR (95% CI)<sup>a</sup> | *P* value |
| --- | --- | --- |
| Natalizumab vs fingolimod | 0.37 (0.14–1.00) | 0.049 |
| Natalizumab vs injectable DMT | 0.15 (0.07–0.31) | <0.001 |
| Fingolimod vs injectable DMT | 0.49 (0.29–0.83) | 0.008 |

<sup>a</sup>Adjusted for prior DMT (naïve vs experienced), count of prebaseline DMTs, and index year.
CI, confidence interval; DMT, disease-modifying therapy; HR, hazard ratio; PS-IPTW, propensity score inverse probability of treatment weighting.

#### Secondary outcomes

The ARRs (95% CI) in patients treated with natalizumab (0.08 [0.05–0.11]) and in patients treated with fingolimod (0.12 [0.08–0.16]) were both significantly lower than the ARR in patients treated with an injectable DMT (0.35 [0.33–0.38]; *P*<0.0001 for both comparisons; Table 3). ARR was not significantly different for natalizumab-treated versus fingolimod-treated patients (RR [95% CI]: natalizumab vs fingolimod, 0.68 [0.41– 1.12] *P*=0.07).

**Table 3.**
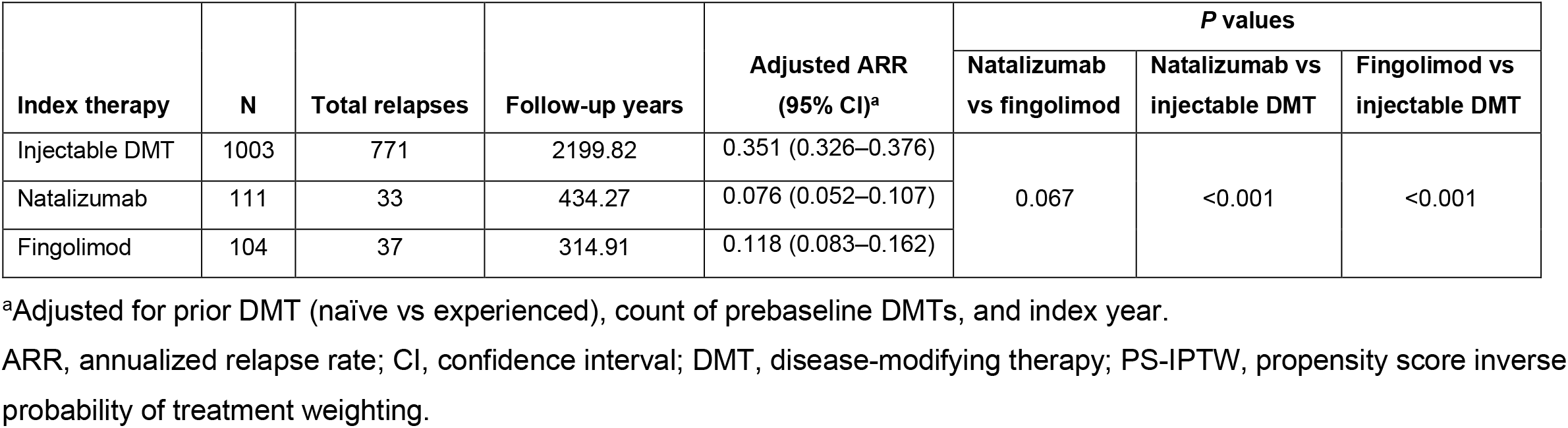
PS-IPTW adjusted annualized relapse rates

The time to index treatment discontinuation was significantly longer in patients treated with natalizumab or fingolimod versus injectable DMT (Figure 2; Table 4), as demonstrated by a lower risk of discontinuing index treatment during follow-up (adjusted HR [95% CI]: natalizumab vs injectable DMT, 0.24 [0.16–0.36]; fingolimod vs injectable DMT, 0.24 [0.15–0.39]; *P*<0.001 for both comparisons). However, the time to discontinuation of index therapy was similar for natalizumab- and fingolimod-treated patients (adjusted HR, 1.00; 95%CI, 0.56–1.78; *P*=0.998).

**Figure 2.**
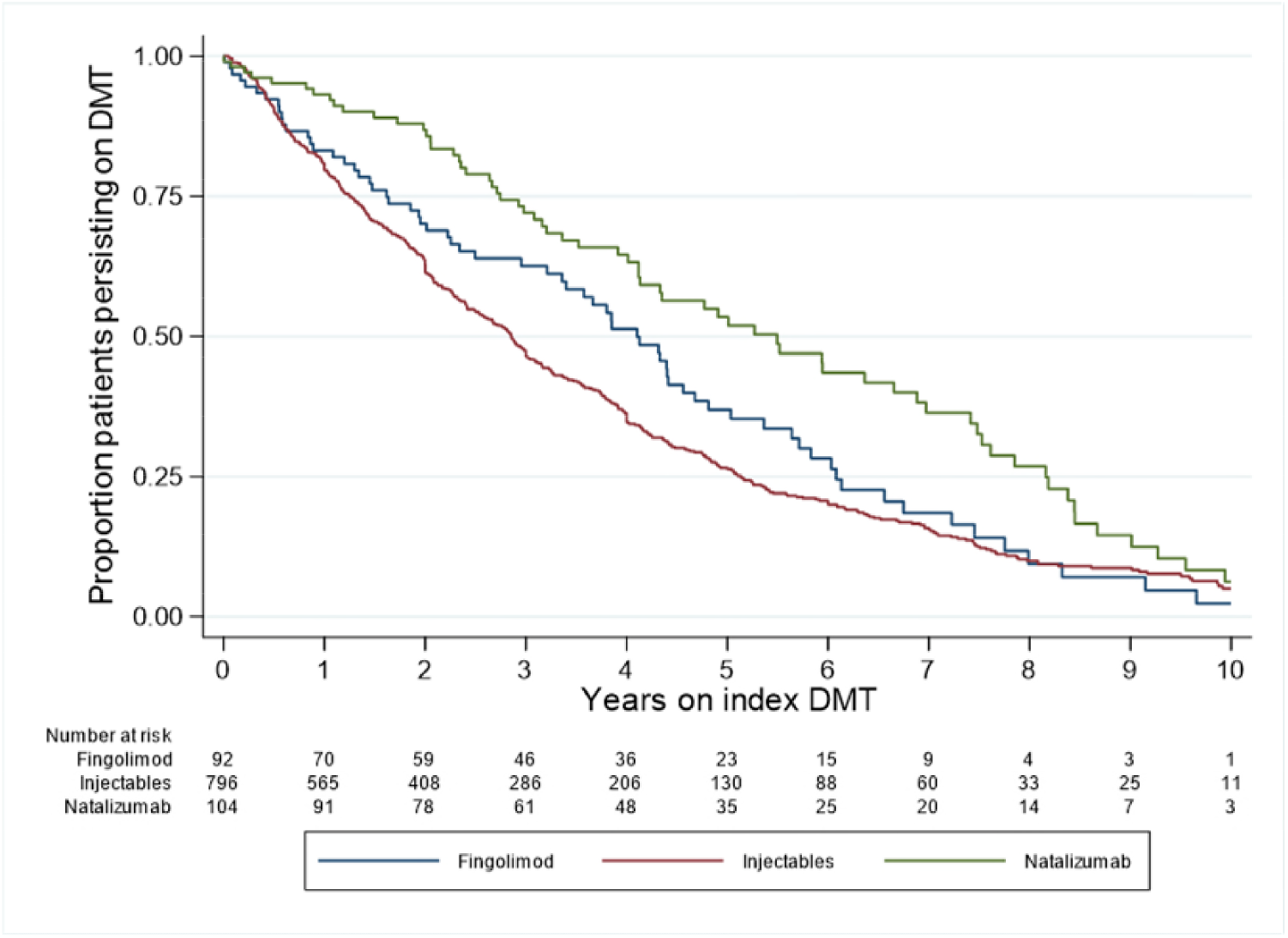
Cumulative probability of remaining on index DMT. DMT, disease-modifying therapy.

**Table 4.** PS-IPTW adjusted hazard ratio estimates of secondary outcomes

| Outcome | PS-IPTW adjusted HR (95% CI)<sup>a</sup> | P value |
| --- | --- | --- |
| **Risk of persisting on index DMT** |  |  |
| Natalizumab vs fingolimod | 1.00 (0.56–1.78) | 0.998 |
| Natalizumab vs injectable DMT | 0.24 (0.16–0.36) | <0.001 |
| Fingolimod vs injectable DMT | 0.24 (0.15–0.39) | <0.001 |
| **Risk of not demonstrating 24-week CDW** |  |  |
| Natalizumab vs fingolimod | 0.84 (0.26–2.75) | 0.782 |
| Natalizumab vs injectable DMT | 2.17 (0.81–5.85) | 0.124 |
| Fingolimod vs injectable DMT | 2.27 (0.73–7.06) | 0.156 |
| **Risk of reaching 24-week CDI** |  |  |
| Natalizumab vs fingolimod | 0.97 (0.52, 1.83) | 0.936 |
| Natalizumab vs injectable DMT | 2.24 (1.33, 3.76) | 0.002 |
| Fingolimod vs injectable DMT | 2.98 (1.70, 5.23) | <0.001 |

<sup>a</sup>Adjusted for prior DMT (naïve vs experienced), count of prebaseline DMT, and index year.
CDI, confirmed disability improvement; CI, confidence interval; CDW, confirmed disability worsening; DMT, disease-modifying therapy; HR, hazard ratio; PS-IPTW, propensity score inverse probability of treatment weighting.

The time to 24-week CDW was not significantly different in any treatment group; however, natalizumab and fingolimod demonstrated a nominally reduced risk of 24-week CDW in comparison with injectable DMTs (Figure 3; Table 4). Adjusted HRs (95% CI) were 2.17 (0.81–5.85; *P*=0.124) for natalizumab versus injectable DMTs and 2.27 (0.73–7.06; *P*=0.156) for fingolimod versus injectable DMTs.

**Figure 3.**
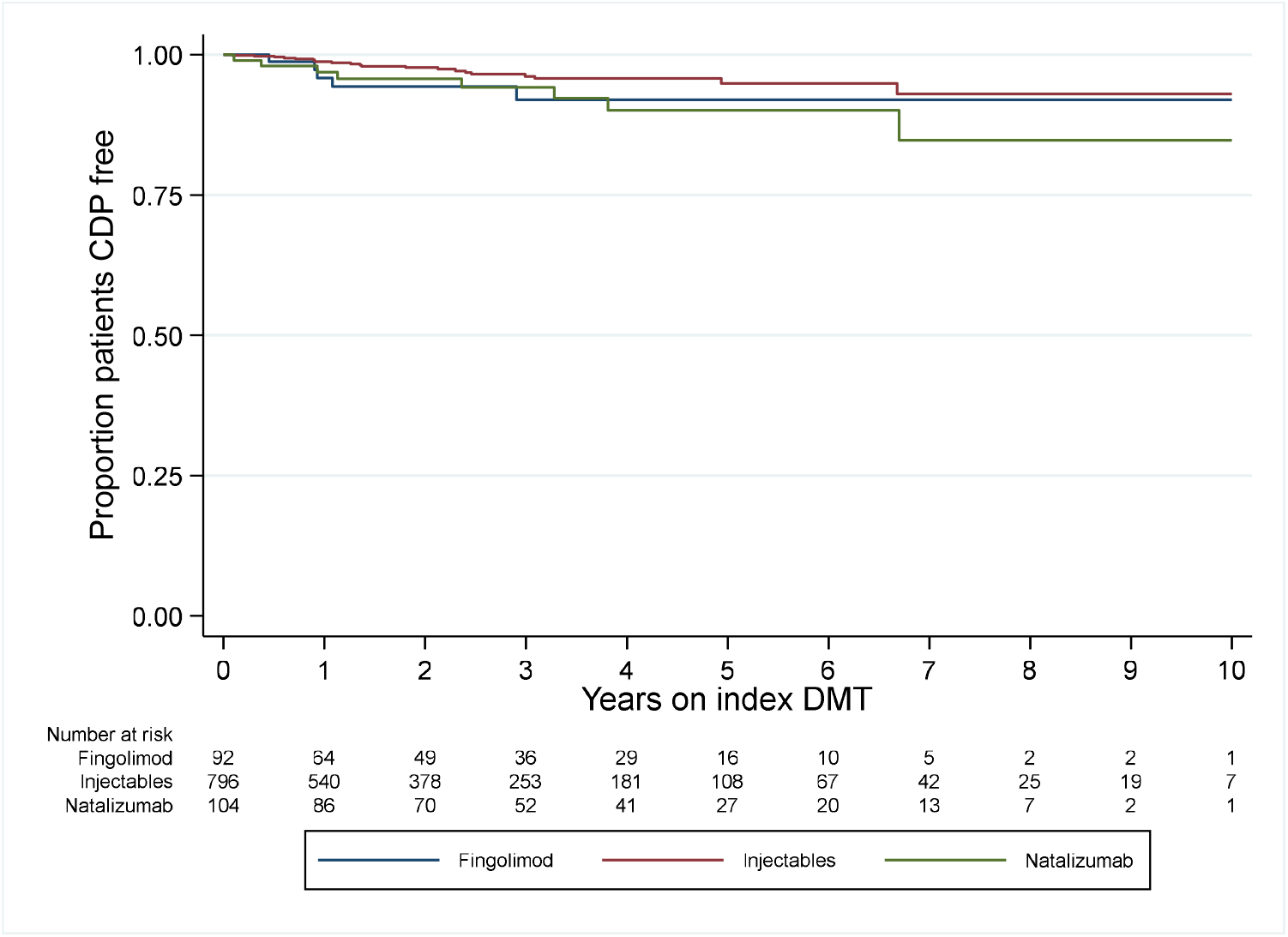
Cumulative probability of remaining free of 24-week CDW for patients treated with natalizumab, fingolimod, or injectable therapies. CDP, confirmed disability progression; CDW, confirmed disability worsening; DMT, disease-modifying therapy.

The cumulative proportion of patients demonstrating 24-week CDI was significantly greater in patients treated with either natalizumab or fingolimod than in patients treated with injectable DMTs (Figure 4; Table 4). Adjusted HRs (95% CI) for time to 24-week CDI were 2.24 (1.33–3.76; p=0.002) for natalizumab versus injectable DMTs and 2.98 (1.70–5.23; p<0.001) for fingolimod versus injectable DMTs.

**Figure 4.**
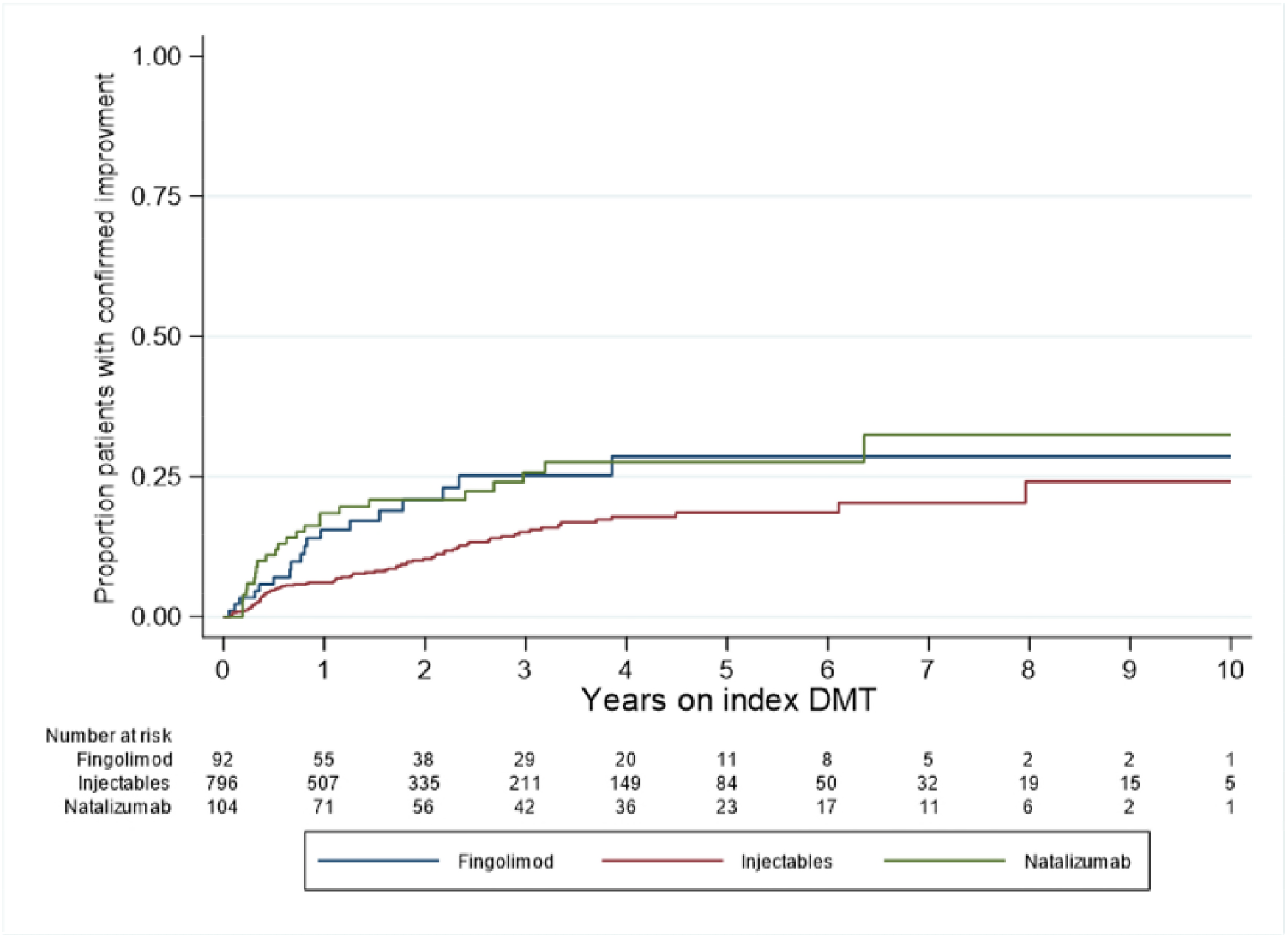
Cumulative probability of reaching 24-week CDI for patients treated with natalizumab, fingolimod, or injectable therapies. CDI, confirmed disability improvement; DMT, disease-modifying therapy.

#### Reasons for index treatment discontinuation

Treatment discontinuations occurred among 709, 76, and 72 patients treated with injectable DMTs, fingolimod, and natalizumab, respectively. Within each treatment group, the most common reported reason for treatment discontinuation was lack of improvement (fingolimod, 29.0%; injectable DMT, 22.6%; natalizumab, 22.2%). The reason for discontinuation was not reported for approximately a third of patients (injectable DMT, 33.9%; natalizumab, 33.3%; fingolimod, 27.6%).

#### Description of monitoring times

Postbaseline visits with an EDSS measurement (mean [standard deviation [SD]) were more common among patients treated with natalizumab (5.9 [7.6]) or fingolimod (4.3 [5.8]) than with an injectable DMT (2.0 [3.9]). However, the intervisit time (mean months between visits with an EDSS evaluation [SD]) was similar between treatment groups (natalizumab, 5.7 [4.9]; fingolimod, 5.0 [3.9]; injectable DMT, 5.3 [4.9]), indicating that visits were conducted at a consistent rate regardless of treatment in patients for whom EDSS was evaluated.

### Sensitivity Analyses

Analyses of 1:1 propensity score–matched patients (n=54 patients in each treatment group) produced similar results to the main IPTW analyses (Supplemental Tables 1–3). The time to first relapse was nominally reduced in matched patients treated with natalizumab or fingolimod in comparison with injectable DMT, as evidenced by a lower risk of relapse during follow-up (HR [95% CI] natalizumab vs injectable DMTs: 0.44 [0.19–1.02], *P*=0.055; fingolimod vs injectable DMTs: 0.48 [0.21–1.11], *P*=0.085). ARR (95% CI) was significantly reduced in propensity score–matched patients treated with natalizumab (0.07 [0.04–0.11]) or fingolimod (0.08 [0.05–0.14]) in comparison with injectable DMT (0.42 [0.27–0.62]) (*P*<0.001 for both comparisons). The time to index treatment discontinuation was significantly longer in matched patients treated with natalizumab or fingolimod than with injectable DMT, as demonstrated by a reduced risk of relapse during follow-up (HR [95% CI]: natalizumab vs injectable DMT, 0.18 [0.10– 0.35]; fingolimod vs injectable DMT, 0.28 [0.15–0.52]; *P*<0.001 for both comparisons). As in the primary analysis, time to 24-week CDW in EDSS was not significantly different in any treatment group. Time to 24-week CDI was also similar to that in the primary analysis, with a significantly increased risk of improvement observed with fingolimod versus injectable DMT (HR, 8.22; 95% CI, 1.07–62.92; *P*=0.043) and nominally increased risk of improvement in patients treated with natalizumab versus injectable DMT (HR, 5.63; 95% CI, 0.72–43.92; *P*=0.099). In pairwise propensity score–matched analyses, no 2 treatments demonstrated significant differences in 24-week CDI.

The results from other sensitivity analyses were also consistent with the primary analysis. A trimmed analysis—which excluded 2 of 111 (1.8%) natalizumab-treated patients, 6 of 104 (5.8%) fingolimod-treated patients, and 297 of 1003 (29.6%) injectable DMT–treated propensity score outlier patients—also generated results consistent with the main analyses (Supplemental Tables 4–6). Results of the ATT-weighted analyses were consistent with the primary analysis regardless of whether patients were weighted to be similar in baseline characteristics to those treated with natalizumab, fingolimod, or injectable DMTs (Supplemental Table 7). Results from the 2010 data cutoff sensitivity analysis were consistent with the 2006 data cutoff, despite the reduction in number of patients treated with natalizumab (n=91) or injectable DMTs (n=724) in this time period (Supplemental Tables 8–10). Finally, results of a sensitivity analysis in 83 of 111 (74.8%) natalizumab-treated patients, 62 of 104 (59.6%) fingolimod-treated patients, and 536 of 1003 (53.4%) injectable DMT–treated patients with baseline MRI data were also consistent with the main primary and secondary analyses (Supplemental Tables 11–13).

## DISCUSSION

Pediatric-onset MS is a rare disease, and patients under the age of 18 are excluded from most randomized MS trials. Due to the relative rarity of the disease and frequent off-label use, randomized trials require many centers and a long recruitment period, and therefore are very expensive and difficult to conduct and complete.^33^ In the absence of randomized clinical trial data, real-world evidence is increasingly used to investigate important clinical questions including MS disease prognosis, predictors of treatment response and long-term outcomes, therapeutic effectiveness, and comparative effectiveness and safety of different DMTs. ^34^

We conducted a 3-way IPTW analysis of real-world clinical data from the MSBase registry to compare the effectiveness of natalizumab, fingolimod and injectable therapies (interferon-beta, glatiramer acetate) in patients with POMS. For the primary study endpoint, patients with POMS who initiated treatment with natalizumab, or who switched to natalizumab from an injectable therapy, showed a greater probability of remaining relapse-free than did those who initiated treatment with fingolimod or who switched to fingolimod from an injectable therapy. Patients treated with either natalizumab or fingolimod had a significantly greater probability of remaining relapse-free than those treated with injectable therapies.

The adjusted secondary study outcomes were generally consistent with the primary outcome or with previous observations. Proportions of patients remaining on index therapy were significantly greater for patients treated with either natalizumab or fingolimod than with an injectable DMT. The time to index therapy discontinuation was essentially the same for natalizumab and fingolimod, and for all 3 groups the most common reason for discontinuation was a lack of improvement.^35^

Time to 24-week CDW was not significantly different among the 3 treatment groups. The lack of a differential treatment effect for natalizumab is consistent with previous studies of natalizumab in adult patients with relapsing-recurring MS in MSBase, and is in part due to the relative rarity of CDP events in treated real-world observational studies .^36^

Time to 24-week CDI was significantly lower in MSBase patients with pediatric MS treated with natalizumab or with fingolimod than in patients treated with an injectable DMT, consistent with previous clinical and real-world observations of patients with AOMS.^37,38^ These observations are also consistent with evidence that natalizumab and fingolimod are effective anti-inflammatory agents, as assessed by reductions in relapse rates, in patients with POMS.^13^

The findings in the IPTW-adjusted patient populations were confirmed in multiple sensitivity analyses. Importantly, the analyses of propensity score–matched patients were generally consistent with the main analyses, as were the analyses of the trimmed IPTW-adjusted treatment groups.

This was a real-world, retrospective cohort study of patients in the MSBase registry and, as such is subject to the limitations typical of real-world analyses generally and of registry-based studies specifically.^34^ Aside from the nonrandomized design inherent to these studies, some residual indication bias can remain, especially in relation to the results of MRI and their association with treatment choice. Head-to-head randomized clinical trials are generally considered to provide the highest-quality evidence for comparative studies of therapeutic agents. For rarer diseases such as POMS, randomized clinical trial data can be limited or unavailable. Real-world studies offer a powerful alternative to randomized clinical trials for these patient populations, and large multicenter networks and patient registries such as MSBase can provide access to more patients than could be enrolled in a randomized clinical trial. Therefore real-world studies can answer practical questions related to treatment and outcomes for rarer patient populations, and real-world data are being increasingly accepted and used by regulatory agencies.^39,40^

When comparing patient cohorts from real-world data sources, patient groups must be balanced to address potential differences in baseline demographic or disease characteristics. Standard propensity score matching is not straightforward with 3 comparator groups, because the target population after matching is not easily defined. Additionally, the algorithms used for matching generally need a larger sample size per treatment arm than was available to us. When weighting with IPTW, variations in baseline characteristics that predict an individual treatment are weighted so as to calculate propensity scores independent of treatment assignment.^41^ Thus, each treatment group may be weighted to mirror baseline characteristics of the overall treated population. Furthermore, IPTW enables sensitivity analyses to compare weighting to baseline characteristics of the full analysis sample and weighting to baseline characteristic patterns present in each individual treatment group.

Even though the 3 treatment groups appeared to be well balanced following IPTW, there is potential for residual bias due to unmeasured covariates. However, the consistent results obtained with the sensitivity analyses argue that such bias, if present, cannot account for the observed results. The similar results of the main and sensitivity analyses are evidence of high internal validity of this study. It is not unreasonable to assume that the pediatric patients in this study who were treated with natalizumab or fingolimod have more severe disease than those treated with injectable DMTs and might therefore be expected to have worse outcomes than those in the injectable group. This was not observed, however, suggesting that indication bias is significantly mitigated.

Overall, the results of this retrospective registry study are consistent with previous studies of natalizumab ^13,22-30^ and fingolimod in patients with POMS, and therefore entirely expected.^21,42-46^ The results are also generally consistent with comparative studies of natalizumab and fingolimod in adult patients with MS, in concordance with the general agreement that pediatric- and adult-onset MS have similar underlying pathophysiology^14^ and that outcomes for patients <18 years are not fundamentally different than those for patients >18 years, although data supporting this point are limited.

Our results suggest that natalizumab and fingolimod have broadly equivalent therapeutic effectiveness in this population of MSBase patients with POMS. However, the consistent results of the analyses of relapse outcomes in these 2 treatment groups suggests natalizumab could be superior to fingolimod on relapse outcomes. This is supported by relapse outcomes from comparative studies of natalizumab and fingolimod in patients with pediatric- or adult-onset MS. ^38,47-49^ It is highly likely that lower relapse rates would also be associated with lower brain and spinal cord lesion accumulation, with potential for differential long-term benefits. In particular, cognitive and productivity outcomes, and risk of secondary progressive MS are potentially reduced with maximal control of the early inflammatory phase of RRMS. MSBase is planning to enhance its data collection for cognition and productivity outcomes in the future. Larger cohorts with longer follow-up will be required to ultimately assess differential effects on protection from secondary progressive MS, as has been demonstrated for early use of high-efficacy therapies in AOMS.

Our findings also demonstrate the usefulness of large MS registries and networks in general, and of the MSBase registry in particular, for comparative effectiveness studies of MS DMTs in rare patient populations that are difficult to study in a randomized setting. These results may also be helpful to healthcare providers and their patients in optimizing relapse control and potentially reducing the risk of persistent long-term disability in POMS.

## Data Availability

Requests for data from qualified researchers should be submitted to the corresponding author

## Acknowledgments

Luke Ward, PhD, of Ashfield MedComms, an Ashfield Health Company (Middletown, CT, USA), assisted with drafting the manuscript, and Celia Nelson of Ashfield MedComms edited and styled the manuscript per journal requirements. A list of contributing members of the

MSBase Study Group is given in supplementary table 14.

## REFERENCES

1. Multiple Sclerosis International Federation. Atlas of MS 3rd edition. Part 2: Clinical management of multiple sclerosis around the world. 2021. https://www.msif.org/wp-content/uploads/2021/05/Atlas-3rd-Edition-clinical-management-report-EN-5-5-21.pdf.

2. McGinley MP, Goldschmidt CH, Rae-Grant AD. Diagnosis and treatment of multiple sclerosis: a review. JAMA. 2021;325(8):765–779.

3. Boiko A, Vorobeychik G, Paty D, Devonshire V, Sadovnick D. Early onset multiple sclerosis: a longitudinal study. Neurology. 2002;59(7):1006–1010.

4. Chitnis T, Glanz B, Jaffin S, Healy B. Demographics of pediatric-onset multiple sclerosis in an MS center population from the Northeastern United States. Mult Scler. 2009;15(5):627–631.

5. Harding KE, Liang K, Cossburn MD, et al. Long-term outcome of paediatric-onset multiple sclerosis: a population-based study. J Neurol Neurosurg Psychiatry. 2013;84(2):141–147.

6. Gorman MP, Healy BC, Polgar-Turcsanyi M, Chitnis T. Increased relapse rate in pediatric-onset compared with adult-onset multiple sclerosis. Arch Neurol. 2009;66(1):54–59.

7. Alroughani R, Boyko A. Pediatric multiple sclerosis: a review. BMC Neurol. 2018;18(1):27.

8. Benson LA, Healy BC, Gorman MP, et al. Elevated relapse rates in pediatric compared to adult MS persist for at least 6 years. Mult Scler Relat Disord. 2014;3(2):186–193.

9. Renoux C, Vukusic S, Mikaeloff Y, et al. Natural history of multiple sclerosis with childhood onset. N Engl J Med. 2007;356(25):2603–2613.

10. Wallach AI, Waltz M, Casper TC, et al. Cognitive processing speed in pediatric-onset multiple sclerosis: Baseline characteristics of impairment and prediction of decline. Mult Scler. 2019;26(14):1938–1947.

11. McKay KA, Friberg E, Razaz N, Alexanderson K, Hillert J. Long-term Socioeconomic Outcomes Associated With Pediatric-Onset Multiple Sclerosis. JAMA Neurol. 2021;78(4):478–482.

12. Baroncini D, Simone M, Iaffaldano P, et al. Risk of Persistent Disability in Patients With Pediatric-Onset Multiple Sclerosis. JAMA Neurol. 2021;78(6):726–735.

13. Jakimovski D, Awan S, Eckert SP, Farooq O, Weinstock-Guttman B. Multiple Sclerosis in Children: Differential Diagnosis, Prognosis, and Disease-Modifying Treatment. CNS Drugs. 2022;36(1):45–59.

14. Bar-Or A, Hintzen RQ, Dale RC, Rostasy K, Bruck W, Chitnis T. Immunopathophysiology of pediatric CNS inflammatory demyelinating diseases. Neurology. 2016;87(9 Suppl 2):S12–19.

15. Waubant E, Ponsonby AL, Pugliatti M, Hanwell H, Mowry EM, Hintzen RQ. Environmental and genetic factors in pediatric inflammatory demyelinating diseases. Neurology. 2016;87(9 Suppl 2):S20–27.

16. Margoni M, Rinaldi F, Perini P, Gallo P. Therapy of Pediatric-Onset Multiple Sclerosis: State of the Art, Challenges, and Opportunities. Front Neurol. 2021;12:676095.

17. Baroncini D, Zaffaroni M, Moiola L, et al. Long-term follow-up of pediatric MS patients starting treatment with injectable first-line agents: A multicentre, Italian, retrospective, observational study. Mult Scler. 2019;25(3):399–407.

18. Krysko KM, Graves J, Rensel M, et al. Use of newer disease-modifying therapies in pediatric multiple sclerosis in the US. Neurology. 2018;91(19):e1778–e1787.

19. Gilenya Prescribing Information. Gilenya® (fingolimod) [prescribing information] In. East Hanover, NJ: Novartis Pharmaceuticals Corp; 2019.

20. Gilenya Summary of Product Characteristics. Gilenya® (fingolimod) [summary of product characteristics]. Nuremberg, Germany: Novartis Pharma GmbH; 2018. In:2018.

21. Chitnis T, Arnold DL, Banwell B, et al. Trial of Fingolimod versus Interferon Beta-1a in Pediatric Multiple Sclerosis. N Engl J Med. 2018;379(11):1017–1027.

22. Pohl D, Rostasy K, Gartner J, Hanefeld F. Treatment of early onset multiple sclerosis with subcutaneous interferon beta-1a. Neurology. 2005;64(5):888–890.

23. Banwell B, Reder AT, Krupp L, et al. Safety and tolerability of interferon beta-1b in pediatric multiple sclerosis. Neurology. 2006;66(4):472–476.

24. Ghezzi A, Amato MP, Capobianco M, et al. Treatment of early-onset multiple sclerosis with intramuscular interferon beta-1a: long-term results. Neurol Sci. 2007;28(3):127–132.

25. Kornek B, Aboul-Enein F, Rostasy K, et al. Natalizumab Therapy for Highly Active Pediatric Multiple Sclerosis. JAMA Neurol. 2013;70(4):469–475.

26. Tenembaum SN, Banwell B, Pohl D, et al. Subcutaneous interferon Beta-1a in pediatric multiple sclerosis: a retrospective study. J Child Neurol. 2013;28(7):849–856.

27. Ghezzi A, Moiola L, Pozzilli C, et al. Natalizumab in the pediatric MS population: results of the Italian registry. BMC Neurol. 2015;15:174.

28. Krysko KM, Graves JS, Rensel M, et al. Real-World Effectiveness of Initial Disease-Modifying Therapies in Pediatric Multiple Sclerosis. Ann Neurol. 2020;88(1):42–55.

29. Palavra F, Figueiroa S, Correia AS, et al. TyPed study: Natalizumab for the treatment of pediatric-onset multiple sclerosis in Portugal. Mult Scler Relat Disord. 2021;51:102865.

30. Simpson A, Mowry EM, Newsome SD. Early Aggressive Treatment Approaches for Multiple Sclerosis. Curr Treat Options Neurol. 2021;23(7):19.

31. Kalincik T, Butzkueven H. The MSBase registry: Informing clinical practice. Mult Scler. 2019;25(14):1828–1834.

32. Butzkueven H, Chapman J, Cristiano E, et al. MSBase: an international, online registry and platform for collaborative outcomes research in multiple sclerosis. Mult Scler. 2006;12(6):769–774.

33. Waubant E, Banwell B, Wassmer E, et al. Clinical trials of disease-modifying agents in pediatric MS: Opportunities, challenges, and recommendations from the IPMSSG. Neurology. 2019;92(22):e2538–e2549.

34. Cohen JA, Trojano M, Mowry EM, et al. Leveraging real-world data to investigate multiple sclerosis disease behavior, prognosis, and treatment. Mult Scler. 2020;26(1):23–37.

35. Schwartz CE, Grover SA, Powell VE, et al. Risk factors for non-adherence to disease-modifying therapy in pediatric multiple sclerosis. Mult Scler. 2018;24(2):175–185.

36. Butzkueven H, Kappos L, Wiendl H, et al. Long-term safety and effectiveness of natalizumab treatment in clinical practice: 10 years of real-world data from the Tysabri Observational Program (TOP). J Neurol Neurosurg Psychiatry. 2020;91:660–668.

37. Kalincik T, Horakova D, Spelman T, et al. Switch to natalizumab versus fingolimod in active relapsing-remitting multiple sclerosis. Ann Neurol. 2015;77(3):425–435.

38. He A, Spelman T, Jokubaitis V, et al. Comparison of switch to fingolimod or interferon beta/glatiramer acetate in active multiple sclerosis. JAMA Neurol. 2015;72(4):405–413.

39. US Food and Drug Administration Center for Drug Evaluation and Research. Considerations for the Use of Real-World Data and Real-World Evidence to Support Regulatory Decision-Making for Drug and Biological Products. Guidance for Industry. In. Silver Spring, MD 2021.

40. European Medicines Agency. A vision for use of real-world evidence in EU medicines regulation. 2021; https://www.ema.europa.eu/en/news/vision-use-real-world-evidence-eu-medicines-regulation. Accessed March 6, 2022.

41. Allan V, Ramagopalan SV, Mardekian J, et al. Propensity score matching and inverse probability of treatment weighting to address confounding by indication in comparative effectiveness research of oral anticoagulants. J Comp Eff Res. 2020;9(9):603–614.

42. Arnold DL, Banwell B, Bar-Or A, et al. Effect of fingolimod on MRI outcomes in patients with paediatric-onset multiple sclerosis: results from the phase 3 PARADIGMS study. J Neurol Neurosurg Psychiatry. 2020;91(5):483–492.

43. Deiva K, Huppke P, Banwell B, et al. Consistent control of disease activity with fingolimod versus IFN β-1a in paediatric-onset multiple sclerosis: further insights from PARADIGMS. J Neurol Neurosurg Psychiatry. 2020;91(1):58–66.

44. Feng J, Rensel M. Review of the Safety, Efficacy and Tolerability of Fingolimod in the Treatment of Pediatric Patients With Relapsing-Remitting Forms of Multiple Sclerosis (RRMS). Pediatric Health Med Ther. 2019;10:141–146.

45. Zaffaroni M. Fingolimod in pediatric-onset multiple sclerosis. Neurol Sci. 2021;42(Suppl 1):1–4.

46. Ziemssen T, Albrecht H, Haas J, et al. Descriptive Analysis of Real-World Data on Fingolimod Long-Term Treatment of Young Adult RRMS Patients. Front Neurol. 2021;12:637107.

47. Cohen M, Mondot L, Bucciarelli F, et al. BEST-MS: A prospective head-to-head comparative study of natalizumab and fingolimod in active relapsing MS. Mult Scler. 2021;27(10):1556–1563.

48. Butzkueven H, Licata S, Jeffery D, et al. Natalizumab versus fingolimod for patients with active relapsing-remitting multiple sclerosis: results from REVEAL, a prospective, randomised head-to-head study. BMJ Open. 2020;10(10):e038861.

49. Guerra T, Caputo F, Orlando B, Paolicelli D, Trojano M, Iaffaldano P. Long-term comparative analysis of no evidence of disease activity (NEDA-3) status between multiple sclerosis patients treated with natalizumab and fingolimod for up to 4 years. Neurol Sci. 2021;42(11):4647–4655.

