## Supplementary Methods for "Comparative effectiveness of natalizumab and fingolimod and injectable therapies in patients with pediatric multiple sclerosis: A registry-based retrospective cohort study"

### Supplemental Materials

**Supplemental Table 1.** Baseline characteristics and standardized differences between treatment groups after propensity score 1:1 matching with replacement

| Characteristic | Natalizumab[a] (n=54) | Fingolimod[b] (n=54) | Injectable DMT[c] (n=54) | Standardized differences | | |
|---|---|---|---|---|---|---|
| | | | | Natalizumab vs fingolimod | Natalizumab vs injectable DMTs | Fingolimod vs injectable DMTs |
| Age, mean (SD), years | 15.93 (0.64) | 15.85 (0.42) | 15.96 (1.77) | 0.121 | 0.113 | −0.042 |
| Female, n (%) | 35 (64.8) | 38 (70.4) | 40 (74.1) | −0.118 | −0.140 | −0.082 |
| Country, n(%) | | | | | | |
| Australia | 5 (9.3) | 10 (18.5) | 9 (16.7) | 0.219 | 0.214 | −0.072 |
| Italy | 3 (5.6) | 2 (3.7) | 2 (3.7) | | | |
| Kuwait | 15 (27.8) | 11 (20.4) | 12 (22.2) | | | |
| Spain | 5 (9.3) | 8 (14.8) | 7 (13.0) | | | |
| Turkey | 3 (5.6) | 1 (1.9) | 2 (3.7) | | | |
| Czech Republic | 2 (3.7) | 2 (3.7) | 2 (3.7) | | | |
| Iran | 0 (0.0) | 0 (0.0) | 0 (0.0) | | | |
| Belgium | 0 (0.0) | 0 (0.0) | 0 (0.0) | | | |
| Canada | 0 (0.0) | 0 (0.0) | 0 (0.0) | | | |
| Other | 18 (33.3) | 15 (27.8) | 16 (29.6) | | | |
| MS duration, mean (SD), years | 1.77 (1.54) | 1.83 (1.93) | 1.54 (1.61) | −0.036 | 0.141 | 0.160 |
| BL EDSS score,[d] median (IQR) | 1.5 (1, 2.5) | 1.5 (1, 2.5) | 1.5 (1, 2.5) | 0.113 | 0.108 | 0.066 |
| Prior DMT use, n (%) | | | | | | |
| Naïve | 30 (55.6) | 36 (66.7) | 34 (63.0) | 0.157 | −0.150 | 0.077 |
| Experienced | 24 (44.4) | 18 (33.3) | 20 (37.0) | | | |

| | | | | | | |
|---|---|---|---|---|---|---|
| Number of prior DMTs, mean (SD) | 0.52 (0.64) | 0.52 (0.93) | 0.48 (0.72) | <0.001 | 0.054 | 0.045 |
| Relapses in year prior to BL, mean (SD) | 1.27 (1.30) | 1.22 (1.30) | 1.26 (0.94) | 0.141 | 0.078 | −0.033 |
| Index year, n (%) | | | | | | |
| 2006–2010 | 7 (13.0) | 1 (1.9) | 2 (3.7) | −0.180 | −0.219 | −0.034 |
| 2011–2015 | 31 (57.4) | 31 (57.1) | 28 (51.9) | | | |
| 2016+ | 16 (29.6) | 22 (40.7) | 24 (44.4) | | | |

[a]Patients did not have prior natalizumab treatment. [b]Patients did not have prior fingolimod treatment. [c]Includes IM or SC interferon beta-1a, SC interferon beta-1b, SC glatiramir acetate, and IM or SC peginterferon beta-1a. [d]Nearest EDSS score within 6 months of BL.

BL, baseline; DMT, disease-modifying therapy; EDSS, Expanded Disability Status Scale; IM, intramuscular; IQR, interquartile range; MS, multiple sclerosis; SC, subcutaneous; SD, standard deviation.

**Supplemental Table 2.** Annualized relapse risk after propensity score 1:1 matching with replacement

| Index therapy | n | Total relapses | Follow-up years | ARR (95% CI) | *P* values | | |
|---|---|---|---|---|---|---|---|
| | | | | | Natalizumab vs fingolimod | Natalizumab vs injectable DMT | Fingolimod vs injectable DMT |
| Injectable DMT | 526 | 25 | 59.7 | 0.419 (0.271–0.618) | 0.629 | <0.0001 | <0.0001 |
| Natalizumab | 83 | 16 | 231.58 | 0.069 (0.039–0.112) | | | |
| Fingolimod | 62 | 15 | 182.54 | 0.082 (0.046–0.136) | | | |

ARR, annualized relapse rate; CI, confidence interval; DMT, disease-modifying therapy.

**Supplemental Table 3.** Kaplan-Meier estimates of relative risk of secondary outcomes after propensity score 1:1 matching with replacement

| Comparison | HR (95% CI) | *P* value |
|---|---|---|
| **Risk of remaining relapse-free** | | |
| Natalizumab vs fingolimod | 0.83 (0.38–1.81) | 0.632 |
| Natalizumab vs injectable DMT | 0.44 (0.19–1.02) | 0.055 |
| Fingolimod vs injectable DMT | 0.48 (0.21–1.11) | 0.085 |
| **Risk of persisting on index DMT** | | |
| Natalizumab vs fingolimod | 0.76 (0.46–1.24) | 0.267 |
| Natalizumab vs injectable DMT | 0.18 (0.10–0.35) | <0.001 |
| Fingolimod vs injectable DMT | 0.28 (0.15–0.52) | <0.001 |
| **Risk of not demonstrating 24-week CDW** | | |
| Natalizumab vs fingolimod | 1.04 (0.23–4.67) | 0.963 |
| Natalizumab vs injectable DMT | 1.42 (0.15–13.25) | 0.757 |
| Fingolimod vs injectable DMT | 1.42 (0.14–13.95) | 0.764 |
| **Risk of reaching 24-week CDI** | | |
| Natalizumab vs fingolimod | 0.73 (0.33–1.61) | 0.439 |
| Natalizumab vs injectable DMT | 5.63 (0.72–43.92) | 0.099 |
| Fingolimod vs injectable DMT | 8.22 (1.07–62.92) | 0.043 |

CDI, confirmed disability improvement; CDW, confirmed disability worsening; CI, confidence interval; DMT, disease-modifying therapy; HR, hazard ratio.

**Supplemental Table 4.** Baseline characteristics and standardized differences between treatment groups after trimming the first quartile of propensity scores

| Characteristic | Natalizumab[a] (n=109) | Fingolimod[b] (n=98) | Injectable DMT[c] (n=706) | Standardized difference, unweighted | | | Standardized difference, weighted | | |
|---|---|---|---|---|---|---|---|---|---|
| | | | | Natalizumab vs fingolimod | Natalizumab vs injectables | Fingolimod vs injectables | Natalizumab vs fingolimod | Natalizumab vs injectables | Fingolimod vs injectables |
| Age, mean (SD), years | 15.87 (2.21) | 15.95 (2.95) | 16.01 (2.05) | 0.000 | 0.000 | −0.025 | 0.000 | 0.004 | −0.007 |
| Female, n (%) | 81 (74.3) | 71 (72.5) | 507 (71.8) | 0.042 | 0.056 | 0.014 | 0.032 | 0.061 | −0.05 |
| Country, n (%) | | | | | | | | | |
| Australia | 14 (12.8) | 21 (21.4) | 63 (8.9) | 0.181 | −0.15 | −0.272 | 0.181 | −0.067 | −0.199 |
| Italy | 10 (9.2) | 2 (2.0) | 65 (9.2) | | | | | | |
| Kuwait | 32 (29.4) | 11 (11.2) | 74 (10.5) | | | | | | |
| Spain | 9 (8.3) | 14 (14.3) | 51 (7.2) | | | | | | |
| Turkey | 3 (2.8) | 23 (23.5) | 149 (21.1) | | | | | | |
| Czech Republic | 5 (4.6) | 2 (2.0) | 38 (5.4) | | | | | | |
| Iran | 0 (0.0) | 0 (0.0) | 0 (0.0) | | | | | | |
| Belgium | 3 (2.8) | 3 (3.1) | 29 (4.1) | | | | | | |
| Canada | 5 (4.6) | 1 (1.0) | 24 (3.4) | | | | | | |
| Other | 28 (25.7) | 21 (21.4) | 213 (30.2) | | | | | | |
| MS duration, mean (SD), years | 1.77 (1.71) | 1.81 (1.16) | 1.35 (1.70) | −0.026 | 0.244 | 0.270 | −0.017 | 0.029 | 0.136 |
| BL EDSS score,[d] median (IQR) | 1.5 (1, 2.5) | 1 (0, 2) | 1.5 (0, 2) | 0.266 | 0.122 | −0.097 | 0.145 | 0.055 | −0.129 |

| | | | | | | | | | |
|---|---|---|---|---|---|---|---|---|---|
| Prior DMT use, n (%) | | | | | | | | | |
| Naïve | 63 (57.8) | 48 (49.0) | 579 (82.0) | 0.177 | 0.546 | −0.738 | 0.134 | −0.252 | −0.4 |
| Experienced | 46 (42.2) | 50 (51.0) | 127 (18.0) | | | | | | |
| Number of prior DMTs, mean (SD) | 0.61 (0.84) | 0.66 (0.81) | 0.20 (0.44) | −0.07 | 0.613 | 0.718 | −0.008 | 0.202 | 0.313 |
| Relapses in year prior to BL, mean (SD) | 1.54 (1.24) | 1.13 (1.16) | 1.14 (1.01) | 0.339 | 0.356 | −0.004 | 0.076 | 0.037 | −0.144 |
| Index year, n (%) | | | | | | | | | |
| 2006–2010 | 21 (19.3) | 1 (1.0) | 245 (34.7) | −0.37 | 0.352 | 0.760 | 0.077 | 0.500 | 0.499 |
| 2011–2015 | 48 (44.0) | 56 (57.1) | 280 (39.7) | | | | | | |
| 2016+ | 40 (36.7) | 41 (41.8) | 181 (25.6) | | | | | | |
| Follow-up time, years[e] | | | | | | | | | |
| Mean (SD) | 3.87 (2.93) | 3.12 (2.77) | 1.98 (2.27) | — | — | — | — | — | — |
| Median (IQR) | 3.19 (1.59, 5.52) | 2.50 (0.80, 4.77) | 1.17 (0.22, 2.96) | — | — | — | — | — | — |

[a]Patients did not have prior natalizumab treatment. [b]Patients did not have prior fingolimod treatment. [c]Includes IM or SC interferon beta-1a, SC interferon beta-1b, SC glatiramir acetate, and IM or SC peginterferon beta-1a. [d]Nearest EDSS score within 6 months of BL. [e]Follow-up time while on index DMT.

BL, baseline; DMT, disease-modifying therapy; EDSS, Expanded Disability Status Scale; IM, intramuscular; IQR, interquartile range; MS, multiple sclerosis; SC, subcutaneous; SD, standard deviation.

**Supplemental table 5.** Annualized relapse risk after trimming the first quartile of propensity scores

| Index therapy | n | Total relapses | Follow-up years | ARR (95% CI) | ***P* values** | | |
|---|---|---|---|---|---|---|---|
| | | | | | **Natalizumab vs fingolimod** | **Natalizumab vs injectable DMT** | **Fingolimod vs injectable DMT** |
| Injectable DMT | 706 | 443 | 1395.82 | 0.317 (0.289–0.348) | 0.297 | <0.001 | <0.001 |
| Natalizumab | 109 | 33 | 421.54 | 0.078 (0.054–0.110) | | | |
| Fingolimod | 98 | 31 | 305.4 | 0.102 (0.069–0.144) | | | |

ARR, annualized relapse rate; CI, confidence interval; DMT, disease-modifying therapy.

**Supplemental table 6.** Kaplan-Meier estimates of relative risk of secondary outcomes after trimming the first quartile of propensity scores

| Comparison | PS-IPTW adjusted HR (95% CI)[a] | *P* value |
|---|---|---|
| **Risk of remaining relapse-free** | | |
| Natalizumab vs fingolimod | 0.38 (0.14–1.01) | 0.053 |
| Natalizumab vs injectable DMT | 0.18 (0.08–0.39) | <0.001 |
| Fingolimod vs injectable DMT | 0.63 (0.36–1.10) | 0.103 |
| **Risk of persisting on index DMT** | | |
| Natalizumab vs fingolimod | 0.93 (0.52–1.67) | 0.821 |
| Natalizumab vs injectable DMT | 0.20 (0.13–0.31) | <0.001 |
| Fingolimod vs injectable DMT | 0.22 (0.13–0.35) | <0.001 |
| **Risk of not demonstrating 24-week CDW** | | |
| Natalizumab vs fingolimod | 0.89 (0.27–2.94) | 0.853 |
| Natalizumab vs injectable DMT | 2.03 (0.67–6.16) | 0.209 |
| Fingolimod vs injectable DMT | 1.95 (0.55–6.94) | 0.301 |
| **Risk of reaching 24-week CDI** | | |
| Natalizumab vs fingolimod | 1.14 (0.61–2.13) | 0.678 |
| Natalizumab vs injectable DMT | 2.47 (1.48–4.12) | 0.001 |
| Fingolimod vs injectable DMT | 2.48 (1.39–4.41) | 0.002 |

[a]Adjusted for prior DMT (naïve vs experienced), count of prebaseline DMT, and index year.

CDI, confirmed disability improvement; CDW, confirmed disability worsening; CI, confidence interval; DMT, disease-modifying therapy; HR, hazard ratio; PS-IPTW, propensity score inverse probability of treatment weighting.

**Supplemental Table 7.** ATT1-, ATT2-, and ATT3-weighted adjusted and unadjusted Kaplan-Meier estimates of relative risk of primary and secondary outcomes

| Comparison | Adjusted[a] | | | | | | Unadjusted | | | | | |
|---|---|---|---|---|---|---|---|---|---|---|---|---|
| | ATT1-weighted | | ATT2-weighted | | ATT3-weighted | | ATT1-weighted | | ATT2-weighted | | ATT3-weighted | |
| | **HR (95% CI)** | ***P* value** | **HR (95% CI)** | ***P* value** | **HR (95% CI)** | ***P* value** | **HR (95% CI)** | ***P* value** | **HR (95% CI)** | ***P* value** | **HR (95% CI)** | ***P* value** |
| **Risk of remaining relapse-free** | | | | | | | | | | | | |
| Natalizumab vs fingolimod | 0.31 (0.12–0.81) | 0.018 | 0.29 (0.11–0.77) | 0.013 | 0.30 (0.11–0.80) | 0.016 | 0.29 (0.12–0.72) | 0.007 | 0.33 (0.13–0.81) | 0.015 | 0.30 (0.12–0.78) | 0.013 |
| Natalizumab vs injectable DMT | 0.14 (0.06–0.29) | <0.001 | 0.14 (0.06–0.29) | <0.001 | 0.15 (0.07–0.31) | <0.001 | 0.11 (0.05–0.23) | <0.001 | 0.09 (0.04–0.19) | <0.001 | 0.11 (0.05–0.23) | <0.001 |
| Fingolimod vs injectable DMT | 0.49 (0.28–0.84) | 0.010 | 0.51 (0.30–0.87) | 0.014 | 0.54 (0.32–0.91) | 0.022 | 0.41 (0.23–0.73) | 0.003 | 0.29 (0.17–0.50) | <0.001 | 0.35 (0.20–0.59) | <0.001 |
| **Risk of persisting on index DMT** | | | | | | | | | | | | |
| Natalizumab vs fingolimod | 0.77 (0.41–1.44) | 0.409 | 0.69 (0.36–1.33) | 0.272 | 0.81 (0.43–1.52) | 0.511 | 0.63 (0.35–1.12) | 0.114 | 0.48 (0.27–0.86) | 0.013 | 0.66 (0.38–1.15) | 0.140 |
| Natalizumab vs injectable DMT | 0.22 (0.15–0.35) | <0.001 | 0.23 (0.15–0.35) | <0.001 | 0.23 (0.15–0.36) | <0.001 | 0.26 (0.17–0.39) | <0.001 | 0.29 (0.20–0.43) | <0.001 | 0.30 (0.20–0.44) | <0.001 |
| Fingolimod vs injectable DMT | 0.29 (0.18–0.46) | <0.001 | 0.29 (0.18–0.46) | <0.001 | 0.29 (0.18–0.47) | <0.001 | 0.40 (0.26–0.62) | <0.001 | 0.46 (0.31–0.68) | <0.001 | 0.46 (0.31–0.68) | <0.001 |

| | | | | | | | | | | | | |
|---|---|---|---|---|---|---|---|---|---|---|---|---|
| **Risk of not demonstrating 24-week CDW** | | | | | | | | | | | | |
| Natalizumab vs fingolimod | 0.69 (0.20–2.45) | 0.568 | 0.65 (0.18–2.32) | 0.510 | 0.71 (0.20–2.51) | 0.591 | 1.02 (0.33–3.15) | 0.970 | 1.04 (0.38–3.20) | 0.945 | 0.93 (0.29–2.96) | 0.896 |
| Natalizumab vs injectable DMT | 2.17 (0.81–5.87) | 0.126 | 2.21 (0.81–6.05) | 0.121 | 2.24 (0.82–6.07) | 0.114 | 2.03 (0.87–4.70) | 0.100 | 2.09 (0.81–5.36) | 0.125 | 2.23 (0.89–5.55) | 0.085 |
| Fingolimod vs injectable DMT | 2.52 (0.79–7.97) | 0.117 | 2.55 (0.81–8.02) | 0.110 | 2.62 (0.83–8.25) | 0.099 | 2.53 (0.83–7.71) | 0.101 | 2.04 (0.68–6.09) | 0.200 | 2.23 (0.75–6.63) | 0.151 |
| **Risk of reaching 24-week CDI** | | | | | | | | | | | | |
| Natalizumab vs fingolimod | 0.86 (0.45–1.64) | 0.645 | 0.79 (0.42–1.51) | 0.476 | 0.90 (0.47–1.72) | 0.751 | 0.91 (0.50–1.68) | 0.773 | 0.96 (0.53–1.77) | 0.906 | 0.91 (0.49–1.70) | 0.763 |
| Natalizumab vs injectable DMT | 2.27 (1.35–3.82) | 0.002 | 2.17 (1.28–3.65) | 0.004 | 2.31 (1.37–3.88) | 0.002 | 1.97 (1.25–3.11) | 0.004 | 1.70 (1.03–2.80) | 0.036 | 1.99 (1.23–3.22) | 0.005 |
| Fingolimod vs injectable DMT | 3.60 (2.03–6.40) | <0.001 | 3.36 (1.90–5.92) | <0.001 | 3.52 (1.99–6.22) | <0.001 | 3.57 (2.00–6.36) | <0.001 | 1.89 (1.08–3.33) | 0.026 | 2.27 (1.30–3.99) | 0.004 |

[a]Adjusted for prior DMT (naïve vs experienced), count of prebaseline DMT, and index year.

ATT1, average treatment effect among patients treated with injectable DMT; ATT2, average treatment effect among patients treated with fingolimod; ATT3, average treatment effect among patients treated with natalizumab; CDI, confirmed disability improvement; CDW, confirmed disability worsening; CI, confidence interval; DMT, disease-modifying therapy; HR, hazard ratio.

**Supplemental Table 8.** Baseline characteristics and standardized differences between treatment groups among patients treated after January 1, 2010

| Characteristic | Natalizumab[a] (n=91) | Fingolimod[b] (n=103) | Injectable DMT[c] (n=724) | Standardized difference, unweighted | | | Standardized difference, weighted | | |
|---|---|---|---|---|---|---|---|---|---|
| | | | | Natalizumab vs fingolimod | Natalizumab vs injectables | Fingolimod vs injectables | Natalizumab vs fingolimod | Natalizumab vs injectables | Fingolimod vs injectables |
| Age, mean (SD), years | 15.74 (2.31) | 16.09 (2.75) | 16.07 (1.98) | −0.135 | −0.152 | 0.006 | 0.018 | −0.026 | −0.043 |
| Female, n (%) | 69 (75.8) | 74 (71.8) | 527 (72.8) | 0.090 | 0.069 | −0.021 | 0.107 | 0.113 | −0.044 |
| Country, n (%) | | | | | | | | | |
| Australia | 12 (13.2) | 20 (19.4) | 35 (4.8) | 0.214 | −0.08 | −0.293 | 0.164 | −0.074 | −0.277 |
| Italy | 6 (6.6) | 2 (1.9) | 42 (5.8) | | | | | | |
| Kuwait | 30 (33.0) | 11 (10.7) | 53 (7.3) | | | | | | |
| Spain | 7 (7.7) | 14 (13.6) | 32 (4.4) | | | | | | |
| Turkey | 3 (3.3) | 25 (24.3) | 217 (30.0) | | | | | | |
| Czech Republic | 3 (3.3) | 2 (1.9) | 18 (2.5) | | | | | | |
| Iran | 0 (0.0) | 0 (0.0) | 33 (4.6) | | | | | | |
| Belgium | 3 (3.3) | 3 (2.9) | 23 (3.2) | | | | | | |
| Canada | 5 (5.5) | 1 (1.0) | 15 (2.1) | | | | | | |
| Other | 22 (24.2) | 25 (24.3) | 256 (35.4) | | | | | | |
| MS duration, mean (SD), years | 1.59 (1.64) | 1.86 (1.69) | 1.30 (1.72) | −0.16 | 0.174 | 0.328 | −0.23 | −0.041 | 0.144 |
| BL EDSS score,[d] median (IQR) | 1.5 (1, 2.5) | 1 (0, 2) | 1.5 (0, 2.5) | 0.161 | 0.067 | −0.088 | 0.167 | 0.121 | −0.138 |

| | | | | | | | | | |
|---|---|---|---|---|---|---|---|---|---|
| Prior DMT use, n (%) | | | | | | | | | |
| Naïve | 57 (62.6) | 47 (45.6) | 631 (87.2) | 0.345 | −0.587 | −0.975 | 0.142 | −0.261 | −0.498 |
| Experienced | 34 (37.4) | 56 (54.4) | 93 (12.9) | | | | | | |
| Number of prior DMTs, mean (SD) | 0.53 (0.82) | 0.70 (0.80) | 0.14 (0.39) | −0.211 | 0.603 | 0.887 | −0.062 | 0.292 | 0.380 |
| Relapses in year prior to BL, mean (SD) | 1.55 (1.18) | 1.13 (1.17) | 1.05 (0.86) | 0.361 | 0.489 | 0.079 | 0.495 | 0.524 | −0.04 |
| Index year, n (%) | | | | | | | | | |
| 2006–2010 | 2 (2.2) | 0 (0.0) | 80 (11.1) | −0.019 | 0.412 | 0.443 | 0.027 | 0.314 | 0.215 |
| 2011–2015 | 49 (53.8) | 59 (57.3) | 433 (59.8) | | | | | | |
| 2016+ | 40 (44.0) | 44 (42.7) | 211 (29.1) | | | | | | |

[a]Patients did not have prior natalizumab treatment. [b]Patients did not have prior fingolimod treatment. [c]Includes IM or SC interferon beta-1a, SC interferon beta-1b, SC glatiramir acetate, and IM or SC peginterferon beta-1a. [d]Nearest EDSS score within 6 months of BL.

BL, baseline; DMT, disease-modifying therapy; EDSS, Expanded Disability Status Scale; IM, intramuscular; IQR, interquartile range; MS, multiple sclerosis; SC, subcutaneous; SD, standard deviation.

**Supplemental Table 9.** Annualized relapse risk among patients treated after January 1, 2010

| Index therapy | N | Total relapses | Follow-up years | ARR (95% CI) | *P* values | | |
|---|---|---|---|---|---|---|---|
| | | | | | Natalizumab vs fingolimod | Natalizumab vs injectable DMT | Fingolimod vs injectable DMT |
| Injectable DMT | 724 | 451 | 1255.51 | 0.359 (0.327–0.394) | 0.1415 | <0.0001 | <0.0001 |
| Natalizumab | 91 | 27 | 316.25 | 0.085 (0.056–0.124) | | | |
| Fingolimod | 103 | 37 | 299.35 | 0.124 (0.087–0.170) | | | |

ARR, annualized relapse rate; CI, confidence interval; DMT, disease-modifying therapy.

**Supplemental Table 10.** PS-IPTW adjusted Kaplan-Meier estimates of relative risk of secondary outcomes among patients treated after January 1, 2010

| **Comparison** | **PS-IPTW adjusted HR (95% CI)[a]** | ***P* value** |
|---|---|---|
| **Risk of remaining relapse-free** | | |
| Natalizumab vs fingolimod | 0.39 (0.15–0.95) | 0.038 |
| Natalizumab vs injectable DMT | 0.16 (0.07–0.36) | <0.001 |
| Fingolimod vs injectable DMT | 0.53 (0.31–0.90) | 0.019 |
| **Risk of persisting on index DMT** | | |
| Natalizumab vs fingolimod | 0.66 (0.39–1.19) | 0.174 |
| Natalizumab vs injectable DMT | 0.19 (0.12–0.31) | <0.001 |
| Fingolimod vs injectable DMT | 0.27 (0.16–0.43) | <0.001 |
| **Risk of not demonstrating 24-week CDW** | | |
| Natalizumab vs fingolimod | 0.74 (0.21–2.62) | 0.636 |
| Natalizumab vs injectable DMT | 1.56 (0.47–5.15) | 0.469 |
| Fingolimod vs injectable DMT | 2.58 (0.82–8.14) | 0.106 |
| **Risk of reaching 24-week CDI** | | |
| Natalizumab vs fingolimod | 0.89 (0.47–1.68) | 0.713 |
| Natalizumab vs injectable DMT | 2.61 (1.50–4.53) | 0.001 |
| Fingolimod vs injectable DMT | 3.22 (1.82–5.68) | <0.001 |

[a]Adjusted for prior DMT (naïve vs experienced), count of prebaseline DMT, and index year.

CDI, confirmed disability improvement; CDW, confirmed disability worsening; CI, confidence interval; DMT, disease-modifying therapy; HR, hazard ratio; PS-IPTW, propensity score inverse probability of treatment weighting.

**Supplemental Table 11.** Baseline characteristics and standardized differences between treatment groups among patients with baseline MRI

| Characteristic | Natalizumab[a] (n=83) | Fingolimod[b] (n=62) | Injectable DMT[c] (n=526) | Standardized difference, unweighted | | | Standardized difference, weighted | | |
|---|---|---|---|---|---|---|---|---|---|
| | | | | Natalizumab vs fingolimod | Natalizumab vs injectables | Fingolimod vs injectables | Natalizumab vs fingolimod | Natalizumab vs injectables | Fingolimod vs injectables |
| Age, mean (SD), years | 15.95 (1.94) | 15.92 (2.76) | 16.16 (1.88) | 0.012 | −0.116 | −0.106 | 0.010 | 0.108 | −0.057 |
| Female, n (%) | 67 (80.7) | 45 (72.6) | 375 (71.3) | 0.192 | 0.221 | 0.029 | 0.117 | 0.105 | −0.086 |
| Country, n(%) | | | | 0.295 | −0.207 | −0.291 | 0.147 | 0.104 | −0.243 |
| Australia | 10 (12.1) | 10 (16.1) | 42 (8.0) | | | | | | |
| Italy | 9 (10.8) | 2 (3.2) | 62 (11.8) | | | | | | |
| Kuwait | 29 (34.9) | 6 (9.7) | 37 (7.0) | | | | | | |
| Spain | 6 (7.2) | 12 (19.4) | 30 (5.7) | | | | | | |
| Turkey | 2 (2.4) | 16 (25.8) | 156 (30.0) | | | | | | |
| Czech Republic | 4 (4.8) | 1 (1.6) | 28 (5.3) | | | | | | |
| Iran | 0 (0.0) | 0 (0.0) | 1 (0.2) | | | | | | |
| Belgium | 0 (0.0) | 0 (0.0) | 0 (0.0) | | | | | | |
| Canada | 0 (0.0) | 0 (0.0) | 0 (0.0) | | | | | | |
| Other | 23 (27.7) | 15 (24.2) | 170 (32.3) | | | | | | |
| MS duration, mean (SD), years | 1.38 (1.32) | 1.61 (1.51) | 1.26 (1.69) | −0.158 | 0.084 | 0.220 | −0.139 | −0.053 | 0.162 |
| BL EDSS score,[d] median (IQR) | 1.5 (1, 2.5) | 1 (0, 2) | 1.5 (1, 2) | 0.472 | 0.275 | −0.191 | 0.173 | 0.175 | −0.149 |

| | | | | | | | | | |
|---|---|---|---|---|---|---|---|---|---|
| Prior DMT use, n (%) | | | | | | | | | |
| Naïve | 52 (62.7) | 30 (48.4) | 449 (85.4) | 0.288 | −0.534 | −0.849 | 0.143 | −0.201 | −0.272 |
| Experienced | 31 (37.4) | 32 (51.6) | 77 (14.6) | | | | | | |
| Number of prior DMTs, mean (SD) | 0.59 (0.90) | 0.65 (0.77) | 0.16 (0.40) | −0.066 | 0.619 | 0.790 | −0.037 | 0.191 | 0.294 |
| Relapses in year prior to BL, mean (SD) | 1.75 (1.24) | 1.32 (1.13) | 1.20 (0.91) | 0.358 | 0.508 | 0.124 | 0.186 | 0.245 | −0.081 |
| Index year, n (%) | | | | | | | | | |
| 2006–2010 | 11 (13.2) | 0 (0.0) | 157 (29.9) | −0.317 | 0.397 | 0.740 | 0.056 | 0.263 | 0.368 |
| 2011–2015 | 38 (45.8) | 33 (53.3) | 218 (41.4) | | | | | | |
| 2016+ | 34 (41.0) | 29 (46.8) | 151 (28.7) | | | | | | |
| MRI lesions, n (%) | | | | | | | | | |
| <9 | 6 (7.2) | 5 (8.1) | 81 (15.4) | −0.651 | −0.169 | 0.401 | −0.322 | −0.150 | 0.155 |
| ≥9 | 43 (51.8) | 25 (40.3) | 187 (35.6) | | | | | | |
| Unknown | 34 (41.0) | 32 (51.6) | 258 (49.1) | | | | | | |
| Presence of Gd+ lesions, n (%) | | | | | | | | | |
| Absent | 9 (10.8) | 6 (9.7) | 114 (21.7) | −0.156 | 0.091 | 0.144 | −0.149 | 0.015 | 0.079 |
| Present | 41 (49.4) | 6 (9.7) | 80 (15.2) | | | | | | |
| Unknown | 33 (39.8) | 50 (80.7) | 332 (63.1) | | | | | | |

[a]Patients did not have prior natalizumab treatment. [b]Patients did not have prior fingolimod treatment. [c]Includes IM or SC interferon beta-1a, SC interferon beta-1b, SC glatiramer acetate, and IM or SC peginterferon beta-1a. [d]Nearest EDSS score within 6 months of BL.

BL, baseline; DMT, disease-modifying therapy; EDSS, Expanded Disability Status Scale; Gd+, gadolinium-enhanced; IM, intramuscular; IQR, interquartile range; MRI, magnetic resonance imaging; MS, multiple sclerosis; SC, subcutaneous; SD, standard deviation.

**Supplemental Table 12.** Annualized relapse risk among patients with baseline MRI

<table>
<tr><th rowspan="2">Index therapy</th><th rowspan="2">N</th><th rowspan="2">Total relapses</th><th rowspan="2">Follow-up years</th><th rowspan="2">ARR (95% CI)</th><th colspan="3">P values</th></tr>
<tr><th>Natalizumab vs fingolimod</th><th>Natalizumab vs injectable DMT</th><th>Fingolimod vs injectable DMT</th></tr>
<tr><td>Injectable DMT</td><td>526</td><td>371</td><td>969.15</td><td>0.382 (0.345–0.424)</td><td rowspan="3">0.2267</td><td rowspan="3"><0.0001</td><td rowspan="3"><0.0001</td></tr>
<tr><td>Natalizumab</td><td>83</td><td>25</td><td>302.38</td><td>0.083 (0.054–0.122)</td></tr>
<tr><td>Fingolimod</td><td>62</td><td>21</td><td>177.92</td><td>0.118 (0.073–0.180)</td></tr>
</table>

ARR, annualized relapse rate; CI, confidence interval; DMT, disease-modifying therapy; MRI, magnetic resonance imaging.

**Supplemental Table 13.** PS-IPTW adjusted Kaplan-Meier estimates of relative risk of secondary outcomes among patients with baseline MRI

| Comparison | PS-IPTW adjusted HR (95% CI)[a] | *P* value |
|---|---|---|
| **Risk of remaining relapse-free** | | |
| Natalizumab vs fingolimod | 0.49 (0.14–1.75) | 0.274 |
| Natalizumab vs injectable DMT | 0.16 (0.06–0.39) | <0.001 |
| Fingolimod vs injectable DMT | 0.59 (0.30–1.18) | 0.137 |
| **Risk of persisting on index DMT** | | |
| Natalizumab vs fingolimod | 0.72 (0.32–1.62) | 0.430 |
| Natalizumab vs injectable DMT | 0.19 (0.11–0.31) | <0.001 |
| Fingolimod vs injectable DMT | 0.23 (0.12–0.44) | <0.001 |
| **Risk of not demonstrating 24-week CDW** | | |
| Natalizumab vs fingolimod | 0.69 (0.15–3.20) | 0.639 |
| Natalizumab vs injectable DMT | 1.74 (0.58–5.16) | 0.321 |
| Fingolimod vs injectable DMT | 2.16 (0.59–7.95) | 0.248 |
| **Risk of reaching 24-week CDI** | | |
| Natalizumab vs fingolimod | 0.74 (0.37–1.46) | 0.377 |
| Natalizumab vs injectable DMT | 1.91 (1.11–3.30) | 0.020 |
| Fingolimod vs injectable DMT | 3.05 (1.63–5.72) | <0.001 |

[a]Adjusted for prior DMT (naïve vs experienced), count of prebaseline DMT, and index year.

CDI, confirmed disability improvement; CDW, confirmed disability worsening; CI, confidence interval; DMT, disease-modifying therapy; HR, hazard ratio; MRI, magnetic resonance imaging; PS-IPTW, propensity score inverse probability of treatment weighting.

**Supplemental Table 14.** MSBase contributors and affiliations

| MSBase Contributor | Affiliation |
|---|---|
| Magd Zakaria | Ain Shams University |
| Vahid Shaygannejad | Isfahan University of Medical Sciences, Isfahan, Iran |
| Sara Eichau | Hospital Universitario Virgen Macarena, Sevilla, Spain |
| Guillermo Izquierdo | Hospital Universitario Virgen Macarena, Sevilla, Spain |
| Nevin Shalaby | Kasr Al Ainy MS research Unit (KAMSU), Cairo, Egypt |
| Riadh Gouider | Razi University Hospital, Tunis, Tunisia |
| Alexandre Prat | CHUM and Universite de Montreal, Montreal, Canada |
| Marc Girard | CHUM and Universite de Montreal, Montreal, Canada |
| Pierre Duquette | CHUM and Universite de Montreal, Montreal, Canada |
| Katherine Buzzard | Box Hill Hospital, Melbourne, Australia |
| Olga Skibina | Box Hill Hospital, Melbourne, Australia |
| Vincent Van Pesch | Cliniques Universitaires Saint-Luc, Brussels, Belgium |
| Guy Laureys | Universitary Hospital Ghent, Ghent, Belgium |
| Liesbeth Van Hijfte | Universitary Hospital Ghent, Ghent, Belgium |
| Ismail Ramadan | Gamal Abd el Naser Hospital, Alexandria, Egypt |
| Talal Al-Harbi | King Fahad Specialist Hospital-Dammam, Khobar, Saudi Arabia |
| Recai Turkoglu | Haydarpasa Numune Training and Research Hospital, Istanbul, Turkey |
| Jeannette Lechner-Scott | University Newcastle, Newcastle, Australia |
| Radek Ampapa | Nemocnice Jihlava, Jihlava, Czech Republic |
| Stella Hughes | Royal Victoria Hospital, Belfast, United Kingdom |
| Julie Prevost | CSSS Saint-Jérôme, Saint-Jerome, Canada |

| | |
|---|---|
| Aysun Soysal | Bakirkoy Education and Research Hospital for Psychiatric and Neurological Diseases, Istanbul, Turkey |
| Yolanda Blanco | Hospital Clinic de Barcelona, Barcelona, Spain |
| Marco Onofrj | University G. d'Annunzio, Chieti, Italy |
| Alessandra Lugaresi | Università di Bologna, Bologna, Italia |
| Gerardo Iuliano | Ospedali Riuniti di Salerno, Salerno, Italy |
| Suzanne Hodgkinson | UNSW, Sydney, Australia |
| Pamela McCombe | University of Queensland, Brisbane, Australia |
| Pamela McCombe | Royal Brisbane and Women's Hospital, Brisbane, Australia |
| Seyed Aidin Sajedi | Golestan University of Medical Sciences, Gogan, Iran |
| Daniele Spitaleri | Azienda Ospedaliera di Rilievo Nazionale San Giuseppe Moscati Avellino, Avellino, Italy |
| Michael Barnett | Brain and Mind Centre, Sydney, Australia |
| Dheeraj Khurana | PGIMER, Chandigarh, India |
| Edgardo Cristiano | Centro de Esclerosis Múltiple de Buenos Aires (CEMBA), Buenos Aires, Argentina |
| Mark Slee | Flinders University, Adelaide, Australia |
| Ricardo Fernandez Bolaños | Hospital Universitario Virgen de Valme, Seville, Spain |
| Jose Luis Sanchez-Menoyo | Hospital de Galdakao-Usansolo, Galdakao, Spain |
| Nikolaos Grigoriadis | AHEPA University Hospital, Thessaloniki, Greece |
| Maria Pia Amato | University of Florence, Florence, Italy |
| Oliver Gerlach | Zuyderland Medical Center, Sittard-Geleen, The Netherlands |
| Abdullah Al-Asmi | Sultan Qaboos University, Al-Khodh, Oman |
| Simu Mihaela | University of Medicine and Pharmacy Victor Babes Timisoara |
| Nevin John | Monash Medical Centre, Melbourne, Australia |
| Yara Fragoso | Universidade Metropolitana de Santos, Santos, Brazil |
| Farouk Talaat | Alexandria universtiy hospital, Alexandria, Egypt |
| Barbara Willekens | Department of Neurology, Antwerp University Hospital, Edegem, Belgium |
| Jose Antonio Cabrera-Gomez | Centro Internacional de Restauracion Neurologica, Havana, Cuba |
| Cristina Ramo-Tello | Hospital Germans Trias i Pujol, Badalona, Spain |
| Allan Kermode | University of Western Australia, Nedlands, Australia |
| Marzena Fabis-Pedrini | University of Western Australia, Nedlands, Australia |
| Bart Van Wijmeersch | Pelt and Hasselt University, Hasselt, Belgium |

| | |
|---|---|
| Jiwon Oh | St. Michael's Hospital, Toronto, Canada |
| Jens Kuhle | Universitatsspital Basel, Basel , Switzerland |
| Claudio Gobbi | Ospedale Civico Lugano, Lugano, Switzerland |
| Tamara Castillo Triviño | Hospital Universitario Donostia and IIS Biodonostia, San Sebastián, Spain |
| Celia Oreja-Guevara | Hospital Clinico San Carlos, Madrid, Spain |
| Angel Perez sempere | Hospital General Universitario de Alicante, Alicante, Spain |
| Eduardo Aguera-Morales | University Hospital Reina Sofia, Cordoba, Spain |
| Bhim Singhal | Bombay Hospital Institute of Medical Sciences, Mumbai, India |
| Elisabetta Cartechini | Azienda Sanitaria Unica Regionale Marche - AV3, Macerata, Italy |
| Claudio Solaro | ASL3 Genovese, Genova, Italy |
| Joyce Pauline Joseph | HOSPITAL KUALA LUMPUR, Kuala Lumpur, Malaysia |
| Ilya Kister | New York University Langone Medical Center, New York, United States |
| Maria Laura Saladino | INEBA - Institute of Neuroscience Buenos Aires, Buenos Aires, Argentina |
| Juan Ignacio Rojas | Hospital Universitario de CEMIC, Buenos Aires, Argentina |
| Justin Garber | Westmead Hospital, Sydney, Australia |
| Anneke van der Walt | The Alfred Hospital, Melbourne, Australia |
| Helmut Butzkueven | Monash University, Melbourne, Australia |
| Richard Macdonell | Austin Health, Melbourne, Australia |
| Eppie Yiu | Royal Children's Hospital Melbourne, Melbourne, Australia |
| Danny Decoo | AZ Alma Ziekenhuis, Sijsele - Damme, Belgium |
| Pierre Grammond | CISSS Chaudière-Appalache, Levis, Canada |
| Thor Petersen | Aarhus University Hospital, Arhus C, Denmark |
| Karim Kotkata | Alexandria University Hospital, Alexandria, Egypt |
| Chris McGuigan | St Vincent's University Hospital, Dublin, Ireland |
| Koen de Gans | Groene Hart Ziekenhuis, Gouda, Netherlands |
| Carlos Vrech | Sanatorio Allende, Cordoba, Argentina |
| Marcos Burgos | Hospital San Bernanrdo, San Bernardo, Argentina |
| Neil Shuey | St Vincents Hospital, Fitzroy, Melbourne, Australia |
| Jennifer Massey | St Vincent's Hospital, Sydney, Australia |
| Deborah Field | Lyell McEwin Hospital, Elizabeth Vale, Australia |

| | |
|---|---|
| Patrice Lalive | Geneva University Hospital, Geneva, Switzerland |
| Shereen Fathi | Maadi MS Center, Cairo, Egypt |
| Sarah Besora | Hospital Universitari MútuaTerrassa, Barcelona, Spain |
| Jamie Campbell | Craigavon Area Hospital, Craigavon, United Kingdom |
| Imre Piroska | Veszprém Megyei Csolnoky Ferenc Kórház zrt., Veszprem, Hungary |
| Csilla Rozsa | Jahn Ferenc Teaching Hospital, Budapest, Hungary |
| Tunde Csepany | University of Debrecen, Debrecen, Hungary |
| Krisztina Kovacs | Péterfy Sandor Hospital, Budapest, Hungary |
| Irene Treviño-Frenk | Instituto Nacional de Ciencias Médicas y Nutrición Salvador Zubirán, Mexico City, Mexico |
| Cees Zwanikken | University Hospital Nijmegen, Nijmegen, Netherlands |
| Jan Schepel | Waikato Hospital, Hamilton, New Zealand |
| Mona AlKahawajah | King Faisal Specialist & Research Centre, Riyadh, Saudi Arabia |
